# Personalised interactive music systems for physical activity and exercise: A systematic review and exploratory meta-analysis

**DOI:** 10.1101/2024.05.28.24308089

**Authors:** Andrew Danso, Tiia Kekäläinen, Friederike Koehler, Keegan Knittle, Patti Nijhuis, Iballa Burunat, Pedro Neto, Anastasios Mavrolampados, William M. Randall, Niels Chr. Hansen, Alessandro Ansani, Timo Rantalainen, Vinoo Alluri, Martin Hartmann, Rebecca S. Schaefer, Johanna Ihalainen, Rebekah Rousi, Kat Agres, Jennifer MacRitchie, Petri Toiviainen, Suvi Saarikallio, Sebastien Chastin, Geoff Luck

## Abstract

Personalised Interactive Music Systems (PIMS) are emerging as promising devices for enhancing physical activity and exercise outcomes. By leveraging real-time data and adaptive technologies, PIMS align musical features, such as tempo and genre with users’ physical activity patterns, including frequency and intensity, enhancing their overall experience. This systematic review and exploratory meta-analysis evaluates the effectiveness of PIMS across physical, psychophysical, and affective domains. Searches across nine databases identified 18 eligible studies, of which six (comprising 17 intervention arms) contained sufficient data for meta-analysis. Random-effects meta-analyses and meta-regression were performed to assess outcomes for physical activity levels, physical exertion, ratings of perceived exertion (RPE), and affective valence. Results showed significant improvements in physical activity levels (*g* = 0.49, CI [0.07, 0.91], *p* = .02, *k* = 4) and affective valence (*g* = 1.68, CI [0.15, 3.20], *p* = .03, *k* = 4), with faster music tempo identified as a significant moderator (*p* = 0.04). No significant effects were observed for RPE (*g* = 0.72, CI [-0.14, 1.59], *p* = .10, *k* = 3) or physical exertion (*g* = 0.79, CI [-0.64, 2.10], *p* = .28, *k* = 5). Substantial heterogeneity and limited study quality indicate the need for more robust, randomised controlled trials to establish the efficacy of PIMS in diverse populations.

## Introduction

Regular physical activity and exercise are fundamental to maintaining and enhancing overall health and well-being. Despite their recognised role in preventing and managing non-communicable diseases such as cardiovascular diseases, cancer, and diabetes, engagement in regular physical activity and exercise remains below the suboptimal level [1]. This deficiency undermines the potential for mental health benefits of physical exercise and its contributions to quality of life [2]. The World Health Organization (WHO) defines physical activity broadly, encompassing all forms of bodily movement generated by skeletal muscles that require energy expenditure, including activities such as walking, sports, and dance (WHO, 2022), whereas exercise has been defined as “physical activity that is planned, structured, repetitive, and purposive, aiming to improve or maintain one or more components of physical fitness” [3:126–127]. However, the broad spectrum of activities categorised as physical activity and exercise often presents challenges in promoting consistent engagement and uptake including individual-level barriers such as motivation and time constraints [4]. Efforts to increase engagement to physical activity and exercise have faced significant challenges, frequently producing inconsistent outcomes as exemplified by interventions such as pedometer-based programs, which show variable effectiveness depending on factors including participant motivation, and engagement [4,5].

### The role of music in enhancing physical activity and exercise

Music’s rhythmic properties have been shown to influence perceptions, ergonomics, and physiological markers associated with physical activity and exercise [6–10]. Available evidence suggests that auditory-motor coupling facilitates predictive synchronisation in physical activity and exercise settings, which can reduce perceived exertion and enhance endurance [11,12]. Additionally, when music aligns with individual preferences, such as through self-selection, it may further increase motivation, improve affective states, induce distraction, and lower perceived effort during physical activities and exercise [7,11].

The integration of music into exercise contexts can be further understood through theoretical frameworks such as the Affective-Reflective Theory (ART) and Dual-Mode Theory. ART emphasises the importance of momentary affective responses—such as pleasure or displeasure—in shaping future exercise behaviours [13,14]. These responses, encapsulated in the construct of “affective valence,” reflect the intrinsic pleasantness or unpleasantness of emotional states that fluctuate based on internal and external stimuli. Conversely, Dual-Mode Theory posits that music’s impact on affective responses is most pronounced at moderate exercise intensities, within a zone of response variability. This zone refers to the range of exercise intensity where affective responses—such as feelings of pleasure or displeasure—are particularly sensitive to individual differences (e.g., fitness level, psychological state) and contextual factors (e.g., music, environment, social setting). In this range, attentional focus and physiological cues mediate affective experiences [15]. While both theories acknowledge the importance of affective responses in exercise, Dual-Mode Theory provides a more nuanced perspective by emphasising intensity-dependent variability and its interaction with individual and contextual factors.

Extending the principles of ART and Dual-Mode Theory, [16] framework highlights how music’s intrinsic properties—such as tempo, rhythm, and harmony—interact with personal and situational moderators, including exercise intensity and individual preferences, to influence affective and behavioural outcomes in exercise. Music operates through three primary mechanisms: regulating affective states, dissociating attention from exertional discomfort, and facilitating temporal prediction and rhythmic synchronisation. These mechanisms are most effective within the zone of response variability, where affective valence dynamically influences exercise engagement [15,17]. Empirical studies consistently demonstrate that personalised music enhances energy efficiency, reduces perceived exertion, and improves adherence by fostering positive affect [17]. Such evidence positions personalised music systems as a key tool for optimising both the immediate and long-term benefits of exercise.

### Personalised Interactive Music Systems (PIMS) in physical activity and exercise

Recently, advances in personalised music technologies have led to the development of Personalised Interactive Music Systems (PIMS), which leverage software, sensors, and computer algorithms to deliver a dynamic, tailored music experience during physical activity and exercise [18,19]. These systems integrate with smartphones and wearable devices to monitor user movements and adjust musical features, such as tempo, style, and timbre, in real time to align with exercise routines, enhancing engagement, and adherence to activity [20,21].

PIMS have been designed for diverse contexts, targeting both intrinsic factors (e.g., motivation and attentional focus) and extrinsic factors (e.g., training guidance). For example, a PIMS, the moBeat system, used real-time interactive music and biophysical feedback to enhance cycling performance by increasing intrinsic motivation and maintaining pace and intensity [12]. Similarly, PIMS interventions for older adults have demonstrated benefits for physical endurance and engagement compared to conventional workout [22]. As mobile interventions incorporating personalisation have been shown to be more effective at enhancing physical activity than non-personalised approaches [23], PIMS hold promise for improving physical activity adherence, reducing the rating of perceived exertion (RPE), and fostering positive affective states during exercise by dynamically tailoring music to individual physiological, affective, and contextual needs [7,12,22].

Due to the relatively recent advancements of PIMS, there is yet limited empirical evidence on their effectiveness across physical activity and exercise-related domains. Such information is essential for informing implementation, replication, and comparative evaluation of interventions aimed at promoting the adherence to physical activity and exercise [24]. While systematic reviews and meta-analyses have explored the general effects of music on physical activity and exercise-related outcomes [6,7,25], these reviews predominantly focused on traditional music listening interventions and did not systematically evaluate the impact of personalised and interactive music systems. By specifically examining PIMS, this review and meta-analysis contribute to understanding how tailored, interactive music interventions influence physical, psychophysical, and affective dimensions of physical activity and exercise engagement, thereby addressing a critical gap in the existing literature.

Therefore, this study combines a systematic review and exploratory meta-analysis to evaluate the effectiveness of PIMS on physical activity and exercise-related outcomes. Specifically, the study synthesises findings on physical activity levels, psychophysical measures (e.g., RPE and physical exertion), and affective outcomes (e.g., affective valence and mood states).

Our main research question is: *How effective are PIMS across physical, psychophysical, and affective outcomes during physical activity and exercise?* This analysis intends to provide early insights into the specificity of PIMS’ effects and identify gaps in the literature that warrant further investigation.

## Methods

This systematic review and meta-analysis was designed based on the Preferred Reporting Items for Systematic Reviews and Meta-Analyses (PRISMA) protocol [26]. The full search strategy can be found in the review registration document ([CRD42023465941](https://www.crd.york.ac.uk/prospero/display_record.php?RecordID=465941)).

### Eligibility criteria

We included: (1) Studies investigating the effect of Personalised Interactive Music Systems (PIMS) on physical activity or exercise, including their effects on motivation, exercise intensity, adherence, or related outcomes; (2) Studies including participants from diverse populations (e.g., sufficiently active and not sufficiently active individuals); (3) Articles in the English language, published from January 2010 to May 2024 in peer-reviewed journals or as published proceedings (conference papers were considered due to the limited number of peer-reviewed studies).

We excluded: (1) Studies from non-peer-reviewed sources, books, dissertations and theses; (2) Articles written in languages other than English; (3) Studies that were not directly related to the effect of PIMS on physical exercise or physical activity.

### Information sources

We searched the following databases: (1) Web of Science, (2) SPORTDiscus, (3) Medline, (4) Embase, (5) ACM Digital library databases, (6) Springer, (7) Google Scholar, (8) IEEE Xplore, and (9) Scopus. The database search was supplemented by a backward snowball search whereby the reference list of all articles was scanned for potential sources. The snowball search continued until no new sources could be identified. The initial inter-rater agreement for the identification of relevant sources was *k* = 0.83, indicating a strong level of agreement among the two individuals performing two independent snowball searches (AD, TK).

### Search strategy

A literature search was performed using terminology related to the effects of PIMS on physical activity and exercise, (“Personali*ed Interactive Music System*” OR “Music Recommendation Algorithm” OR “Music Recommendation System*” OR “Streaming” OR “MP3” OR “Digital Music” AND (“Physical Activity” OR “Exercise” OR “Recovery” OR “Recuperation” OR “Sedentary Behav*” OR “Physical Inactivity”).

### Selection process and data collection process

The citations of all retrieved articles were imported into Zotero, where duplicates were systematically identified and removed. Subsequently, two authors (AD, TK) independently screened the titles and abstracts of the studies using ASReview [27] and Rayyan [28].

Articles that could not be definitively excluded based on the title or abstract underwent full-text retrieval for further evaluation. The full-text articles were then independently assessed for inclusion by the same two authors (AD, TK). Disagreements at any stage were resolved through discussion, with a third author consulted to achieve consensus when necessary.

### Data extraction

The studies’ information was extracted to a spreadsheet, including study characteristics, such as the type of PIMS, the study design, PIMS measurement, and the target behaviour of the PIMS (Target Physical Activity or Exercise; Table 2).

### Pre-registration deviations

Where available, quantitative data suitable for meta-analysis were extracted. This was done for the pre-registered outcome of physical activity level, as well as for affective valence, RPE, and physical exertion, which were not pre-registered as outcomes. The decision to extract data on these additional outcomes was taken because of the close relationships between these variables and physical activity and exercise participation, their prevalence as outcomes in the included studies, and the limited number of studies reporting data on physical activity and exercise behaviour. In cases where effect sizes could not be readily calculated based on the published articles, their authors (*n* = 2) were contacted at least twice for additional data, resulting in the provision of calculations for five additional effect sizes.

### Operationalisation of terms

This review operationalises four key terms central to physical activity and exercise research. Physical Activity Level is defined by the quantified volume (e.g., daily activity counts, weekly minutes), intensity (e.g., metabolic equivalent MET, %VO2R), and compliance (e.g., adherence to heart rate zones or regimens) [29,30] of physical activity. Affective Valence refers to the pleasure-displeasure dimension of emotional responses during or after physical activity, assessed using self-report scales such as the Feeling Scale (FS) [31] and the “good vs. bad mood” subscale of the Multidimensional Mood Questionnaire (MDMQ), [32]. These measures capture subjective ratings of positivity or negativity without incorporating arousal [14,33,34]. Physical Exertion encompasses physiological (e.g., heart rate), biomechanical (e.g., stride length), and perceptual demands, providing a comprehensive assessment of effort [35]. These constructs serve as the primary outcomes of interest in this review. The constructs are summarised in Table 1 and described further in S1 Appendix.

**Table 1.** Operationalisation of terms.

| Term | Definition | Operational Metrics | References |
| --- | --- | --- | --- |
| Physical Activity Level | Encompasses the volume, intensity, and compliance with physical activity recommendations or exercise regimens. | **Volume:** Total activity counts per day (TAC/d) via accelerometer, mean weekly minutes. Intensity: Absolute intensity using METs, relative intensity as % VO2R.<br>**Compliance:** Adherence to recommendations or regimens via changes in volume, device usage, or adherence to heart rate zones. | [29,30] |
| Physical Exertion | Effort exerted to perform physical activity, involving physiological, biomechanical, and perceptual demands. | **Physiological:** Heart rate as an indicator of cardiovascular response. **Biomechanical:** Stride length and pace for activities such as running and walking. **Perceptual:** Integration of physiological and biomechanical cues to assess overall effort. | [35] |
| RPE (Rating of Perceived Exertion) | Subjective numerical value reflecting perceived effort during physical activity, integrating sensory cues and physiological sensations. | **Scale:** Borg RPE Scale for aerobic activities (cycling, running). **Category-Ratio Scale:** Borg CR-10 to measure perceived exertion or other sensations. **Responses:** Local sensations (muscles, skin, joints) and central factors (cardio-pulmonary system). | [7,36,37] |
| Affective Valence | The subjective feeling of pleasure or displeasure experienced during or after physical activity. It is independent of perceived exertion and reflects emotional responses to exercise, influenced by individual, contextual, and social factors. | Affective valence is measured using self-report scales, such as:<br>**Scale:**<br>Feeling Scale (FS): Bipolar scale from +5 (very good) to -5 (very bad).<br>PANAS (Positive and Negative Affect Schedule): Assesses positive and negative emotions.<br>Multidimensional Mood Questionnaire (MDMQ): Evaluates mood during exercise using subscales for “good vs. bad mood,” “calmness vs. agitation,” and “alertness vs. tiredness.” Only the “good vs. bad mood” subscale aligns with the pleasure-displeasure dimension of affect.<br>**Context:**<br>Measurement occurs during or immediately after exercise. | [14,31–33] |

### Study risk of bias assessment

The quality of the studies was assessed by two authors (AD, TK) using the Joanna Briggs Institute critical appraisal checklist, including tools for Quasi-Experimental Appraisal, Qualitative Research Appraisal, and the Revised Checklist for Randomised Controlled Trials [JBI, 38] (Fig 2).

### Data synthesis and analysis methods

We conducted a narrative synthesis, categorising studies into two groups based on design: (1) experimental studies, including randomised, quasi-experimental, pilot, and within-subject designs, and (2) proof-of-concept and user-testing studies. This classification enabled the identification of trends within and across these categories.

For experimental studies, we examined outcomes related to physical activity levels, physical exertion, RPE, and affective valence. Proof-of-concept and user-testing studies were analysed for their focus on PIMS design features and effectiveness, including synchronisation, user engagement, and personalisation.

Our synthesis followed the methodological framework of [39], facilitating systematic comparisons across study groups. Trends and variations in PIMS outcomes were interpreted through subgroup analyses, accounting for methodological rigour and study design. We also considered sample characteristics, including demographic variability (e.g., age, fitness level, and population type) and sample size heterogeneity (ranging from *N* = 10 to *N* = 150). Limitations arising from study heterogeneity were explicitly addressed to provide transparency regarding factors affecting generalisability.

Hedges’ *g* effect sizes and standard errors were calculated using [40] tool and analysed in the JASP environment (version 2024). Random-effects meta-analyses, using the inverse variance weighting method [DerSimonian & Laird method, 41] were conducted for physical activity level, physical exertion, RPE and affective valence. These outcomes were selected based on the pre-registration criterion: “Meta-analyses will be performed when at least three studies provide data sufficient for effect size calculation.” For inclusion in the meta-analysis, physical activity outcomes analysed included behaviours such as walking, running, weight training, cycling, housework, and gardening, while studies focusing on non-physical activity outcomes (e.g., subjective feasibility of PIMS) were excluded. Six studies (comprising 17 intervention arms) met this criterion, while outcomes with insufficient data were excluded. Heterogeneity was assessed using the *I²* statistic (relative proportion of variability attributable to heterogeneity), *τ²* statistic (absolute variance), and Cochran’s *Q* statistic (a formal test of homogeneity). To address the dispersion of effects across studies, the prediction interval was calculated, as it provides insights into the range of effects expected in future comparable studies, beyond the mean effect size [42]. A sensitivity analysis was conducted to evaluate publication bias by examining the relationship between standard errors and effect size estimates. Following Sterne and colleagues [43,44] funnel plots were produced to assess asymmetry, while forest plots were used to summarise the data. Data and syntax files for these analyses are available in S1 file.

An exploratory meta-regression analysis was conducted to investigate potential moderators contributing to variability in the effectiveness of PIMS on physical activity level, affect, RPE, and physical exertion. Candidate moderators were selected based on their theoretical relevance to physical activity and exercise research: study size, participant age, exercise intensity, and music tempo. Music tempo was categorised into tempo ranges to standardize data across studies with differing methodologies, reflecting its established influence on motivational and psychophysical responses [45]. Exercise intensity was classified using MET guidelines to enhance comparability [46]. Participant age and study size were included to address population-level and methodological variability, respectively. Due to the small number of studies included in the meta-analyses, the meta-regression encompassed all outcomes of interest, with a focus on generating hypotheses for future research.

Specifically, within the meta-regression analysis, music tempo was categorised into three distinct groups based on beats per minute (BPM): Slow (60–90 BPM), coded as 1; Medium (91–130 BPM), coded as 2; and Fast (131+ BPM), coded as 3. When studies reported variable tempos, the average BPM or the dominant tempo range was used for classification. Exercise intensity was categorised using a three-level scale aligned with MET guidelines [46]: Low (<3 METs), coded as 1; Moderate (3–6.9 METs), coded as 2; and High (≥7 METs), coded as 3. For studies that did not explicitly report METs, intensity was inferred from descriptions of the exercise type or target heart rate zones. Participant age was handled as follows: for studies reporting mean age directly, the provided value was used. In studies reporting age ranges, the midpoint of the range was used as an estimate. If group-specific mean ages were available, a weighted average was calculated based on group sizes to derive an overall mean age for the study.

## Results

### Study selection

**\*All records excluded by ASReview** [27,28].

**Fig 1.**
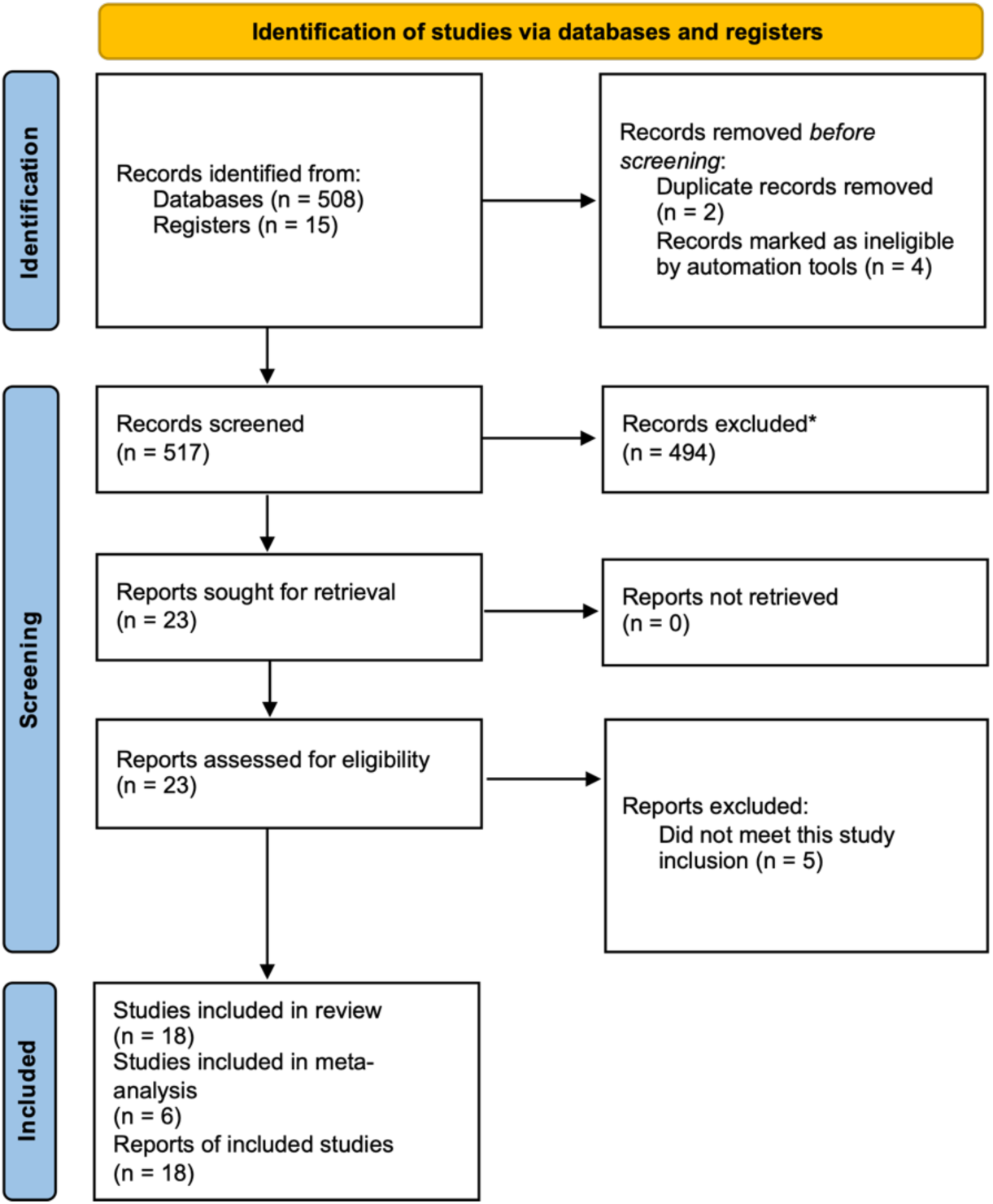
PRISMA information flow describing the screening process. A total of 523 articles were identified through the initial strategic search using the specified keywords. During the screening process, three articles were excluded as duplicates, while four additional articles were identified as ineligible based on the inclusion criteria. After screening titles and abstracts, 494 articles were excluded for not meeting the inclusion criteria. Subsequently, 23 full-text articles were assessed for eligibility. Of these, five articles were excluded because they did not evaluate the desired effect or outcome. In total, 18 articles were eligible to be included in this review study (Fig 1).

### Study characteristics

The study characteristics (Table 2) encompass a diverse range of studies conducted across various countries, including Canada, Spain, Germany, Taiwan, Singapore, Denmark, Finland, Belgium, Switzerland, the Czech Republic, the Netherlands, Norway, and locations not specified. These studies, conducted between 2010 and 2024, provide a broad age range among participants, with some studies focusing on specific groups such as the elderly, patients with cardiovascular disease, students, non-athletes and office workers. The PIMS used in these studies vary in their design and objectives, ranging from personalised music audio-playlists [47–49] to interactive music systems linked to fitness devices [22,34]. These systems are utilised in different settings and for various purposes, ranging from synchronising movement during physical activity and exercise, to enhancing the experience of physical activity and exercise.

**Table 2.** Characteristics of included studies.

| Reference | Country | Age, yrs | Sample Size, N | Population | Type of PIMS | Study Design | PIMS Measurement | Target Behaviour/Target Physical Activity or Exercise | Physical Activity Results | *g* [95% CI] of PIMS on outcomes of interest |
| --- | --- | --- | --- | --- | --- | --- | --- | --- | --- | --- |
| [50] | Canada | 47.3 – 79.2 <sup>a</sup> | 34 | Patients with cardiovascular disease | Personalised music audio-playlists | Randomised Experimental Design | Tri-axial accelerometer | Adherence | Improved PA volumes ($p < 0.001$) | $g = 0.06$ [-1.04, 0.92] for Physical Activity Level (No RAS)<br>$g = 0.51$ [-0.47, 1.49] for Physical Activity Level (RAS) |
| [47] | Spain | N/A | N/A | N/A | Personalised music recommendation system | Proof of Concept | Sensors <sup>b</sup> | Motivation/performance enhancement | N/A | N/A |
| [51]* | Germany | N/A | 1 | Elderly participant | Music feedback for rehabilitation | Proof of Concept | Accelerometer | Rehabilitation | N/A | N/A |
| [52]* | Taiwan | 21.56 ± 1.04 | 10 female and 26 male | Participants from the National Yang Ming Chiao Tung University | Exercise system for middle-distance running | Experimental | Smartphone's built-in tri-axial accelerometer | Adapting music selection to user's pace during walking | N/A | $g = -0.73$ [-1.40, -0.06] for Physical Exertion<br>$g = 1.63$ [0.88, 2.39] for RPE<br>$g = 2.17$ [1.34, 2.99] for Affective Valence |
| [53] | Taiwan | N/A | N/A | N/A | Music Assisted Run Trainer (MART) | Proof of Concept | Tri-axial accelerometer | Physiological, perceptual, affective responses | N/A | N/A |
| [48]* | Singapore | N/A | 60 | Students | A music recommendation system | Within-subjects Crossover Design | Music recommendation ratings | Motivation | N/A | N/A |
| [34] | N/A | N/A | 45 | Non-athletes, non-body builders, non-musicians | Jymmin® – Sensor attached to fitness devices to provide musical feedback | Experimental Design | Movement sensor <sup>c</sup> | Workout | N/A | $g = 0.96$ [0.35, 1.56] for Affective Valence |
| [54]* | N/A | N/A | 27 | N/A | Runner's Jukebox (RJ) – music tempo matching the user's pace during exercise | User Testing Design | Smartphone app to recognise user pace/adjust music tempo | Walking/running pace monitor <sup>d</sup> | N/A | $g = 2.76$ [1.63, 3.89] for Physical Exertion (Fixed BPM)<br>$g = 2.29$ [1.24, 3.34] for Physical Exertion (Pace-Matched)<br>$g = -0.74$ [-1.61, 0.13] for Physical Exertion (Random) |
| [55]* | Denmark | N/A | N/A | N/A | Music clips with dynamic BPM ranging from 110 – 170 | Proof of Concept | Sensors <sup>e</sup> | Cycling | N/A | N/A |
| [56] | Switzerland | 18–45 | 7 females, 8 males | Cyclists | SoundBike – Musical sonification to improve spontaneous synchronisation of cyclists | Experimental | Sensors | Cycling | Enhanced cyclist synchronisation to external music | N/A |
| [57] | Finland | N/A | 2 | Elderly participants | Processing accelerometry data to create musical sonifications of physical activity | Proof of Concept | Sonification of PA data <sup>f</sup> | Awareness of PA | N/A | N/A |
| [58]* | Belgium | N/A | 33 | Participants from public event | DSaT algorithm for music selection and real-time adaptation <sup>g</sup> | Pilot Study | Tri-axial accelerometer | Synchronisation to beat of music | Majority (56.79%) synchronised steps with music | N/A |
| [59] | Belgium, Czech Republic | 21.9 ± 12.9<br>20.2 ± 0.8<br>21.2 ± 1.7<br>23 ± 3 SD | 82 male, 68 female<br>56 male, 44 female<br>12 female<br>6 female, 4 male | N/A | Synchronise music with the participant's movements | Case Study | Recordings of footfalls & music alignment strategies<sup>h</sup> | Synchronisation to beat of music | Improved entrainment | N/A |
| [49]* | N/A | N/A | N/A | N/A | Context-aware recommender system (CAMRS) | Mixed-method Design<sup>i</sup> | Automatic learning algorithm | Motivate users to complete PA | N/A | N/A |
| [22] | Germany | 70.6 (SD ± 3.9) | 11 females<br>5 males | Non-physically active | Jymmin® – Sensor attached to fitness devices to provide musical feedback | Within-subjects Design | Movement sensor<sup>j</sup> | Strength-endurance exercises | N/A | $g = 0.73$ [0.00, 1.46] for Physical Activity Level<br>$g = 0.20$ [-0.27, 0.67] for RPE<br>$g = 0.09$ [-0.47, 0.64] for Affective Valence |
| [60]* | Netherlands | 18 – 25 | 24 | Office workers | Smart cushion providing musical feedback | Within-Subjects Design | Movement sensor-pad | Posture changes | No effect breaking sedentary behaviour | N/A |
| [61] | Norway | N/A | 3 – 6<sup>k</sup> | Seniors with early-stage Alzheimer's disease | Interactive music system | Qualitative Research Design | Sensor-pad | Stimulate/motivate PA | N/A | N/A |
| [12] | Netherlands | 23 – 51 | 26 | Philips employees | MoBeat – Interactive music system | Within-Subject Experiment | Cadence sensor, heart-rate | Motivation | N/A | $g = 0.54$ [-0.22, 1.30] for Physical Activity Level<br>$g = 0.54$ [-0.22, 1.30] for Physical Exertion<br>$g = 0.43$ [-0.32, 1.19] for RPE<br>$g = 3.79$ [2.52, 5.06] for Affective Valence |

*Notes:*
\* = Conference papers.
<sup>a</sup> = Lowest lower bound: 47.3 years (from the second subgroup). Highest upper bound: 79.2 years (from the first subgroup). The estimated entire age range for all three groups combined would be from approximately 47.3 years to 79.2 years.
<sup>b</sup> = Galvanic Skin Response, oxygen saturation sensor, pulse sensor.
<sup>c</sup> = Jymmin® – The movement of the sensor-equipped fitness device is mapped to musical parameters, creating an acoustic feedback signal.
<sup>d</sup> = Swings Per Minute (SWPM).
<sup>e</sup> = Monitor cycling pace and heart rate, influencing audio feedback (soundscape sounds) in real-time.
<sup>f</sup> = Accelerometry data.
<sup>g</sup> = Dynamic Song and Tempo (DSaT).
<sup>h</sup> = The methodology involved recording footfalls and various music alignment strategies to synchronise music with participants' walking or running movements.
<sup>i</sup> = Includes elements of a proof of concept design and an experimental design.
<sup>j</sup> = Jymmin® – The movement of the sensor-equipped fitness device is mapped to musical parameters, creating an acoustic feedback signal.
<sup>k</sup> = Exact numbers are not specified, but a mention of a group size of 3 to 6 participants.

### Reported outcome measures

A variety of outcome measures were reported across studies to explore the effects of PIMS on physical activity and exercise-related behaviours. The outcome measures included assessments of physical activity levels, such as accelerometer-based metrics and adherence to specific heart rate zones, as well as psychological and perceptual outcomes such as mood (measured through tools such as the MDMQ and FS) and intrinsic motivation (measured via the Intrinsic Motivation Inventory, IMI). The rating of perceived exertion (RPE) was frequently captured using the Borg CR-10 scale [36,37]. Table 3 presents this information. Further information on these outcome measurements can be found in S2 Appendix.

**Table 3.**
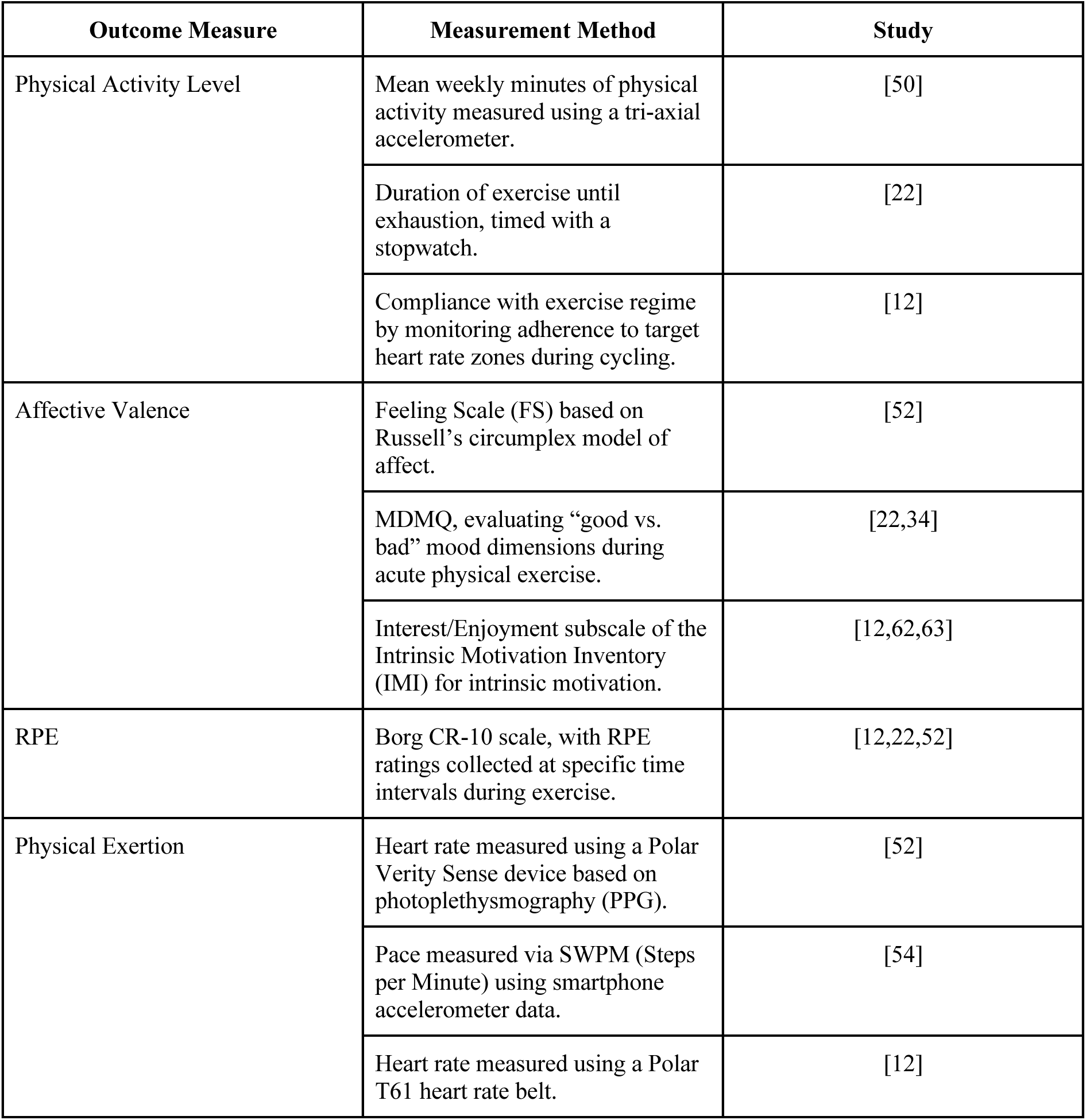
Outcome measures as reported in the studies.

Studies utilised diverse technologies and protocols to assess PIMS’ effects on physical activity and exercise behaviours. Reported technologies included accelerometers, heart rate monitors, and systems such as Jymmin®, which integrate real-time musical feedback with gym equipment. Analytical methods, such as ANOVA and MANOVA, were used to evaluate outcomes, with specific systems adapting music based on cadence, heart rate, and intensity. Table 4 presents a detailed overview of these technologies and protocols.

**Table 4.** Technologies and Analysis Protocols of PIMS.

| Type of PIMS | PIMS Description | Data Analysis Protocol | Reference |
| --- | --- | --- | --- |
| Accelerometer | Music synchronisation with step cadence | N/A | [54] |
| Heart Rate Monitor | Music tempo adjustments based on physiological data | ANOVA, MANOVA | [52] |
| Jymmin® | Music feedback system | ANOVA, MANOVA, Wilcoxon Signed-Ranks Test | [22,34] |
| Magnet and Heart Rate Sensors | Magnet sensors detect RPM, paired with heart rate to optimise cycling rhythms | N/A | [55] |
| MoBeat | Music feedback system | ANOVA | [12] |
| Musical Sonification Systems | Converts movement data into sound to enhance engagement and differentiate activity patterns | One proportion z-test | [57] |
| Musical Sonification (Custom Pedals) | Pedals with load sensors and microcontroller adjust musical feedback | ANOVA, Friedman Test, Pairwise comparisons | [56] |
| PAM® (Personal Activity Monitor) | N/A | Generalised Linear Modelling | [50] |
| Three-axis Accelerometer (Smartphone) | Adjusts music tempo to synchronise with user pace using Swings Per Minute (SWPM) | N/A | [54] |
| Tri-axial Accelerometer | Uses accelerometer and heart rate to adjust music tempo for maintaining target heart rate during cardio-training | N/A | [53] |

### Risk of bias in studies

Following the assessment of the study quality using the JBI critical appraisal checklist tools, the nine criteria were adapted to the five risk of bias domains found in the [64] R package for risk-of-bias assessments (robvis). This assessment tool tests the risk of bias resulting from the randomisation process (D1), deviations from intended intervention (D2), missing outcome data (D3), measurement of the outcome (D4), and selection of the reported result (D5). Each domain is assessed with a judgement scale indicating a high risk of bias (red cross), some concerns (yellow circle), low risk of bias (green plus) and No Information (blue question mark) (cf. Fig 2).

**Fig 2.**
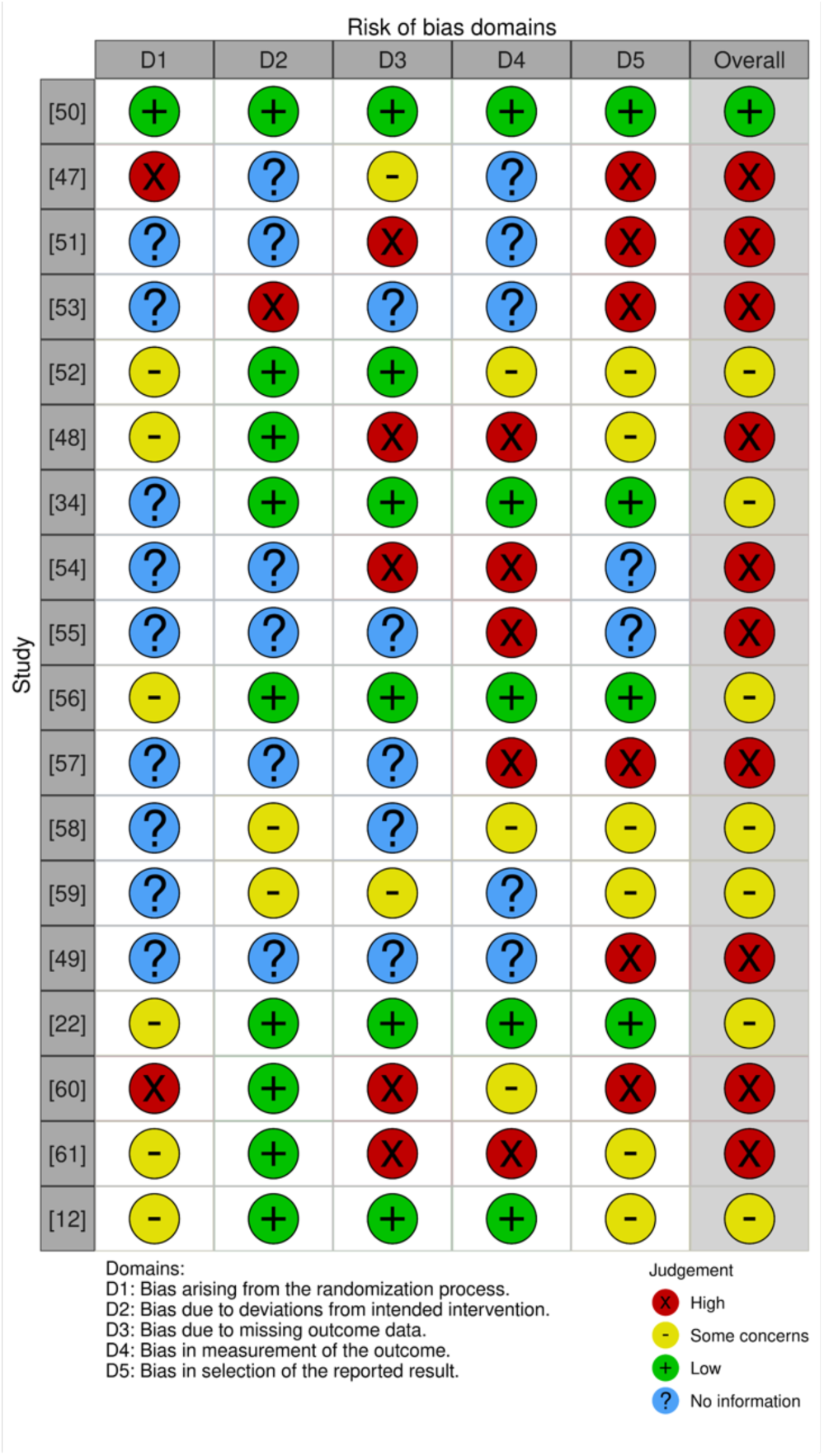
Evaluation of risk of bias in the included studies, categorised across five domains from D1 to D5 [cf.62]. An overall bias risk assessment for each study is also provided, conservatively summarising the findings across all five domains. We included all 18 studies in the review regardless of their overall risk of bias rating (see Fig 2, column ‘Overall’). The overall risk of bias rating for each study was assigned conservatively, reflecting the highest risk level present across any of the five domains (D1– D5). For example, if one domain was judged to have a high risk of bias, the overall rating for that study was classified as high risk. Of the 18 studies, one randomised experimental design study [50] was rated for low risk of bias. Seven studies received a moderate (some concerns) rating of risk of bias, and 10 were rated for a high risk of bias.

### PIMS used in experimental studies

PIMS were explored in experimental studies for their influence on physical, psychophysical, and affective exercise-related outcomes. Several studies focused on synchronisation and auditory-motor coupling. [59] examined beat synchronisation using the D-Jogger adaptive music player. They found that initiating music in phase synchrony significantly enhanced consistent sensorimotor patterns, while strategies relying on tempo adjustments alone were less effective. [56] provided detailed analyses of synchronisation strength using SoundBike, where musical sonification significantly increased pedal cadence synchronisation with external music. Similarly, [54] found significant increases in step frequency (SWPM) when music tempo aligned with user pace, enhancing consistency and efficiency of the activity. [22,34] reported on the Jymmin® system’s role in improving mood and exercise duration. Notably, [34] observed mood enhancements in younger adults, while [22] noted prolonged exercise durations in elderly participants despite no significant mood changes. This may be potentially due to age-related differences in energy pacing. [61] explored a tempo-responsive system for Alzheimer’s patients, observing improved synchronisation and engagement. Additionally, [12] reported the moBeat system maintained exercise compliance while enhancing intrinsic motivation and attentional dissociation from discomfort. Sample sizes varied (*N* = 10 to 150), with participants aged 18–79 across diverse demographics. Detailed descriptions of PIMS used in these studies can be found in S1 Table.

### PIMS used in proof of concept and user testing studies

Proof-of-concept and user-testing studies used PIMS to adapt music or audio feedback based on real-time physical activity and exercise-related data (e.g., heart rate, oxygen saturation, and galvanic skin response, GSR), with a focus on music recommendation systems and synchronisation features. [47] tested DJ-Running, which integrates environmental (GPS), and GSR data to provide personalised music recommendations using algorithms such as Artificial Neural Networks (ANNs). [49] developed a context-aware recommender system using smartphone sensors to adjust music based on exercise intensity, providing evidence for preliminary efficacy in low-concentration activities (e.g., low-to-moderate-intensity activities that require minimal concentration such as walking).

Two synchronisation-based systems were included: [53] Music Assisted Run Trainer (MART), which adjusts music tempo to heart rate or step frequency; and [51] Music Feedback Exercise (MFE) system, which synchronises music with movement intensity through advanced audio processing. For example, as exercise intensity increases, additional layers of musical elements such as rhythm guitar, bass, or drums are progressively added to the audio track. [57] introduced musical sonification, converting movement data into music for users to identify different physical activity patterns. [55] examined ecological soundscapes to influence cycling behaviour. Soundscapes were, for example, dynamically altered based on users’ cycling speed and heart rate. [58] reported optimal movement entrainment at ∼120 BPM using D-Jogger but noted disruptions during song transitions. [48] reinforcement learning-based system found improved user satisfaction and fewer track rejections, while [61] found tempo-responsive music systems beneficial for older adults with Alzheimer’s. Median sample size was *n* = 6, with limited demographic data reducing generalisability. Details on these systems can be found in S2 Table.

### Meta-analyses

A single overall meta-analysis of the studies was not achievable due to heterogeneity across datasets and outcomes [65]. Instead, the outcomes were reported separately based on their focus. The reported outcomes distinguished between (1) physical activity levels, (2) physical exertion, (3) ratings of perceived exertion (RPE), and (4) affective valence.

### Results for physical activity level

The overall effect size is 0.49 with a 95% CI of 0.07 to 0.91, and a *p*-value of .02 (*k* = 4, *n* = 76). This indicates that the results are statistically significant, supporting the effectiveness of PIMS in improving outcomes relating to physical activity level (Fig 3). The random-effects model indicates low heterogeneity (*Q* = 1.65, *p* = .65, *I²* = 0%, Tau = 0.00) between the studies, suggesting it to be negligible. The calculated 95% prediction interval for the true effect size is -0.43 to 1.41, indicating that while the average effect is positive, the range of potential true effects across future studies could include negative or larger positive outcomes.

**Fig 3.**
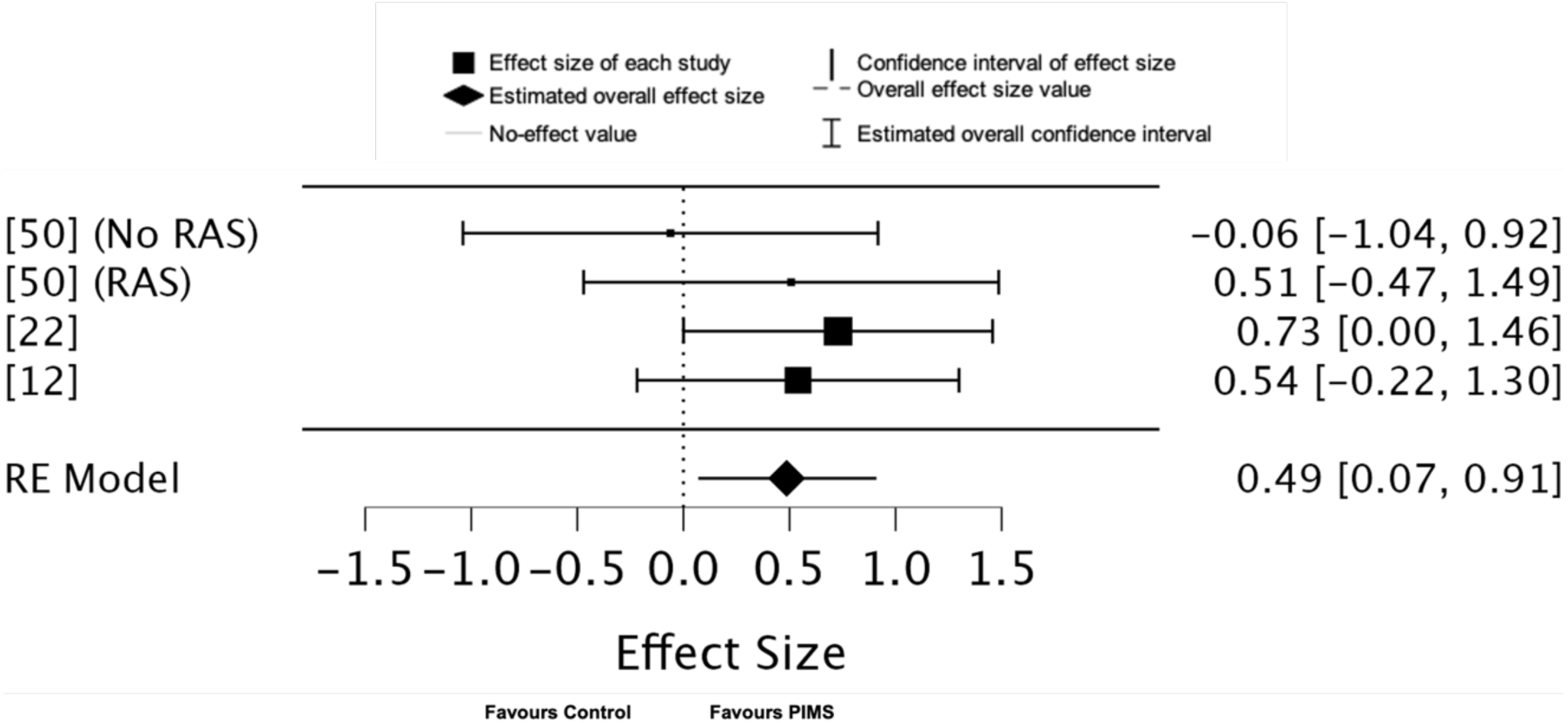
Forest plot of effect sizes for physical activity level outcomes associated with PIMS.

### Results for physical exertion

The overall effect size is 0.79 with a 95% CI of -0.64 to 2.21, and a *p*-value of .28 (*k* = 5, *n* = 142), indicating that the results are not statistically significant and do not support the effectiveness of PIMS in improving physical exertion outcomes (Fig 4). The random-effects model indicates high heterogeneity (*Q* = 46.96, *p* = .00, *I²* = 93%, Tau = 2.44) between the studies. The calculated 95% prediction interval for the true effect size is -4.69 to 6.27, indicating the potential for considerable variation in the effects of PIMS on physical exertion across future studies.

**Fig 4.**
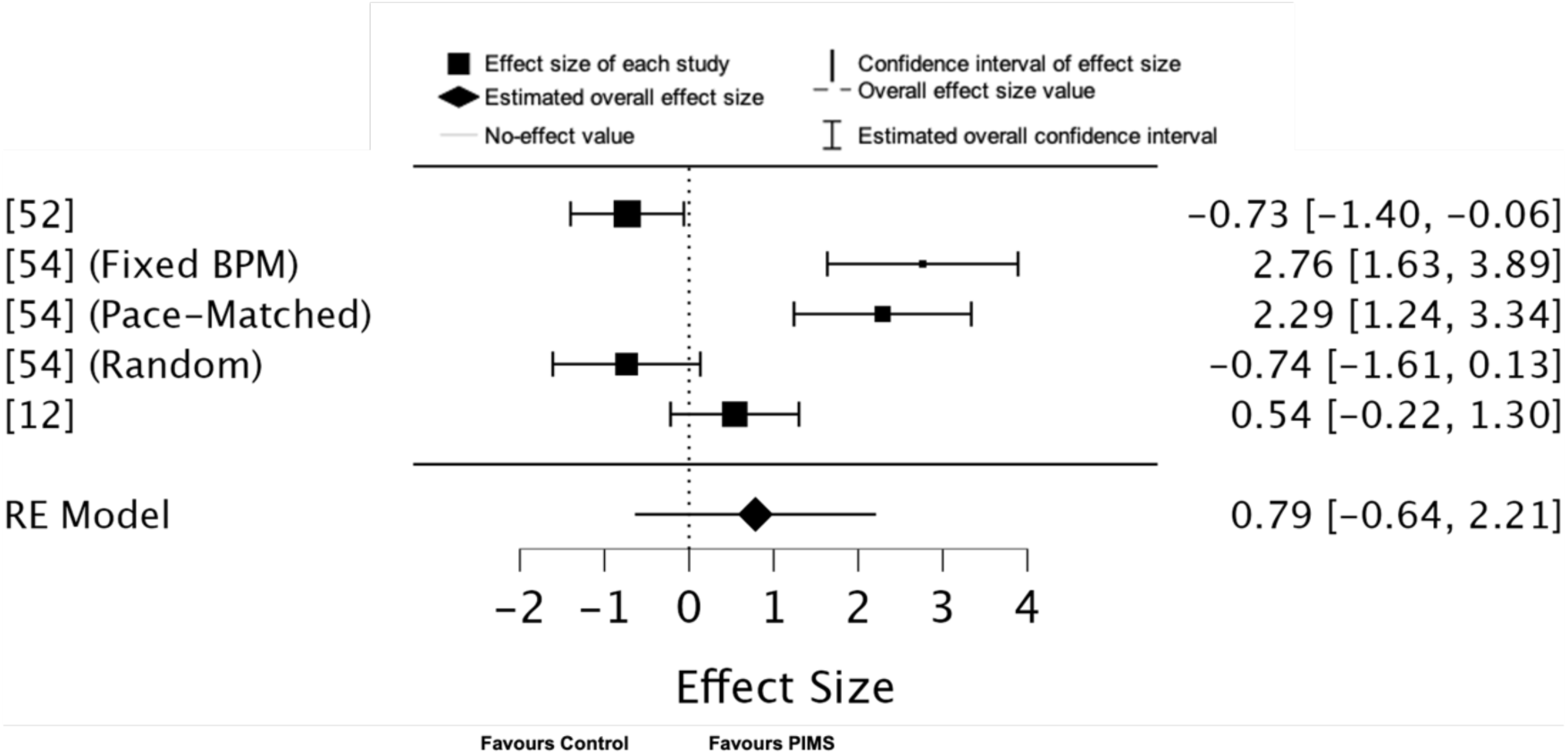
Forest plot of effect sizes for physical exertion outcomes associated with PIMS.

### Results for ratings of perceived exertion (RPE)

The overall effect size is 0.72 with a 95% CI of -0.14 to 1.59, and a *p*-value of .10 (*k* = 3, *n* = 77), indicating that the results are not statistically significant and do not conclusively support the effectiveness of PIMS in improving RPE outcomes (Fig 5). The random-effects model indicates substantial heterogeneity (*Q* = 10.24, *p* = .01, *I²* = 81%, Tau = 0.47) between the studies. The calculated 95% prediction interval for the true effect size is -9.64 to 11.08, reflecting the significant variability in potential outcomes across future studies.

**Fig 5.**
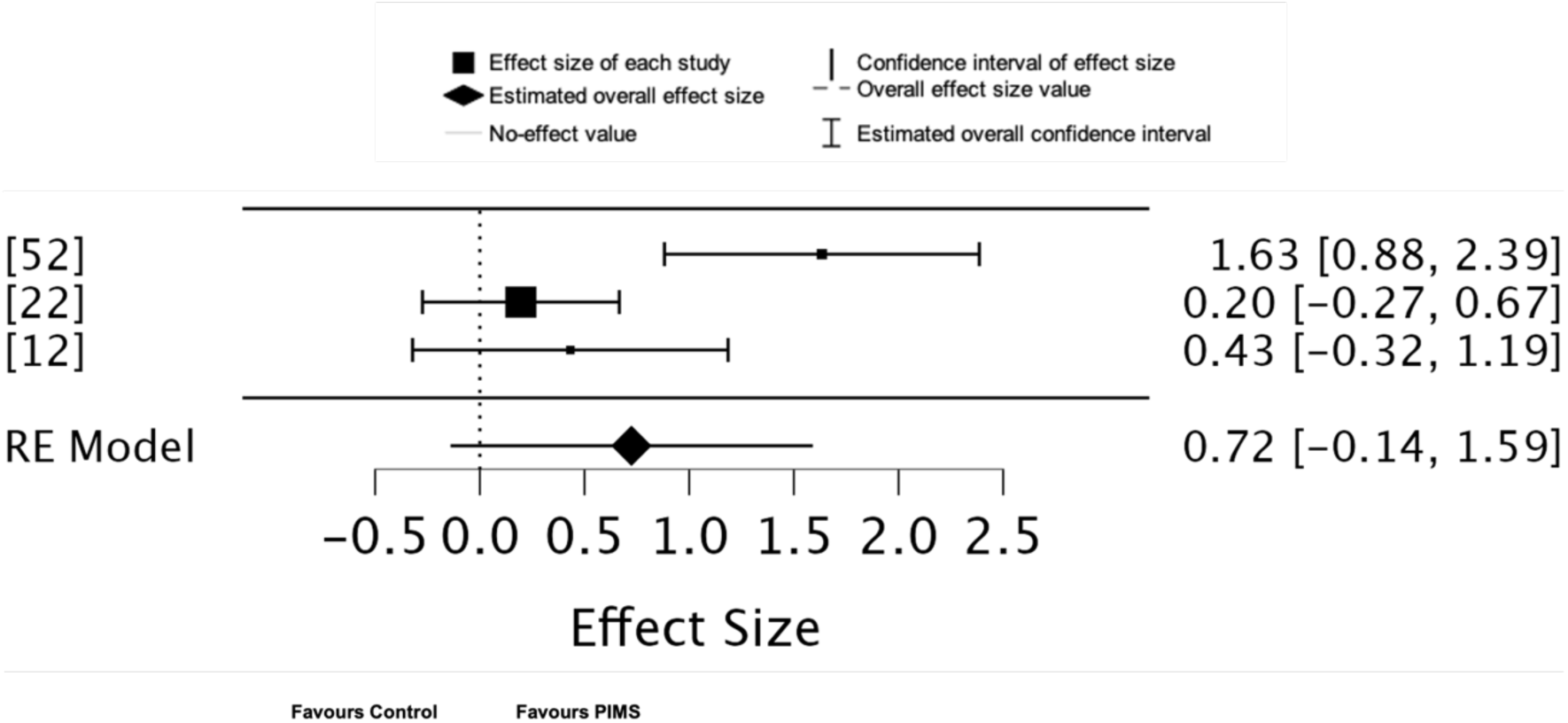
Forest plot of effect sizes for Ratings of Perceived Exertion (RPE) outcomes associated with PIMS.

### Results for affective valence

The overall effect size is 1.68 with a 95% CI of 0.15 to 3.20, and a *p*-value of .03 (*k* = 4, *n* = 122), indicating that the results are statistically significant and thus consistent with the effectiveness of PIMS in improving affective valence outcomes (Fig 6). The random-effects model indicates substantial heterogeneity (*Q* = 36.69, *p* = .00, *I²* = 94%, Tau = 2.24) between the studies. The calculated 95% prediction interval for the true effect size is -5.58 to 8.94, highlighting significant variability in potential outcomes across future studies.

**Fig 6.**
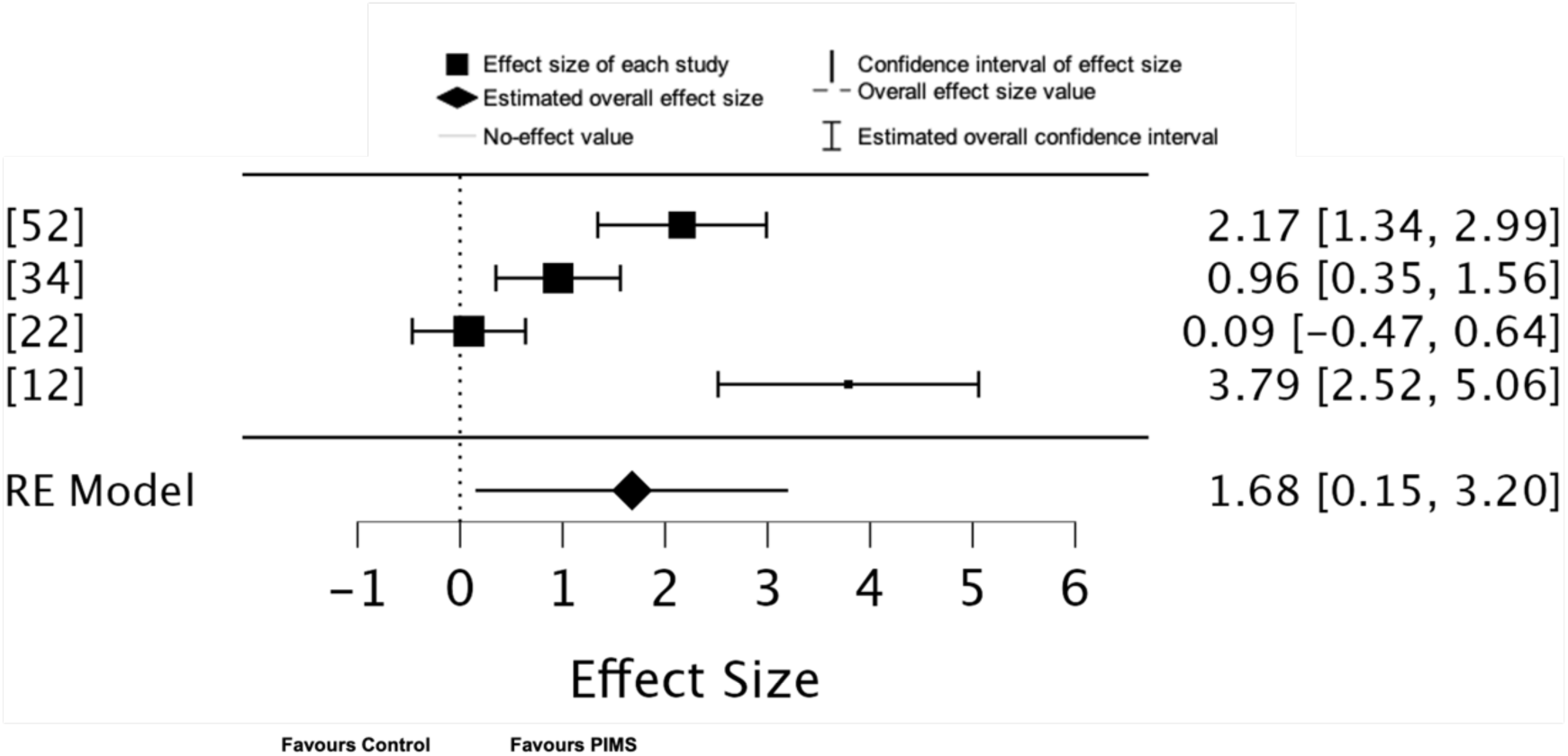
Forest plot of effect sizes for affective valence outcomes associated with PIMS.

### Meta-regression analysis

Heterogeneity was identified in the meta-analyses, prompting the use of meta-regression analysis to explore potential moderators of effect sizes. Music tempi showed a statistically significant positive association with effect sizes (*p* = .044), suggesting that faster tempi may have a significant effect across the outcomes of interest. None of the other predictors, including participant age, exercise intensity, or sample size, demonstrated a significant effect on effect sizes (see Table 5). The overall meta-regression model was not statistically significant (*p* = .189), and substantial heterogeneity remained unexplained (*Q* = 76.782, *p* < .001, *I²* = 87.77%, Tau = 1.049). This indicates that other, unexplored factors likely contribute to the variability in outcomes. Given the inclusion of all outcomes of interest in this analysis, the potential for residual variability and unaccounted-for heterogeneity is high.

**Table 5.** A summary of the meta-regression analysis.

| | Estimate | Standard Error | $z$ | $p$ |
| --- | --- | --- | --- | --- |
| Intercept | 1.229 | 3.208 | 0.383 | .702 |
| Age | -0.036 | 0.043 | -0.822 | .411 |
| Music Tempo | 0.623 | 0.309 | 2.015 | .044 |
| Exercise Intensity | 0.207 | 1.360 | 0.152 | .879 |
| Sample Size | -0.026 | 0.067 | -0.381 | .703 |

*Note.* Wald test.

### Publication bias analysis (Egger’s test)

Egger’s test [66] indicated non-significant asymmetry for physical activity level (*z* = -0.968, *p* = .333), significant asymmetry for physical exertion (*z* = 2.927, *p* = .003), non-significant asymmetry for RPE (*z* = 0.832, *p* = .405), and significant asymmetry for affective valence (*z* = 4.961, *p* < .001) (Fig 7). Because of potential publication bias, the summary effect sizes for physical exertion and affective valence outcomes may thus be slightly inflated.

**Fig 7.**
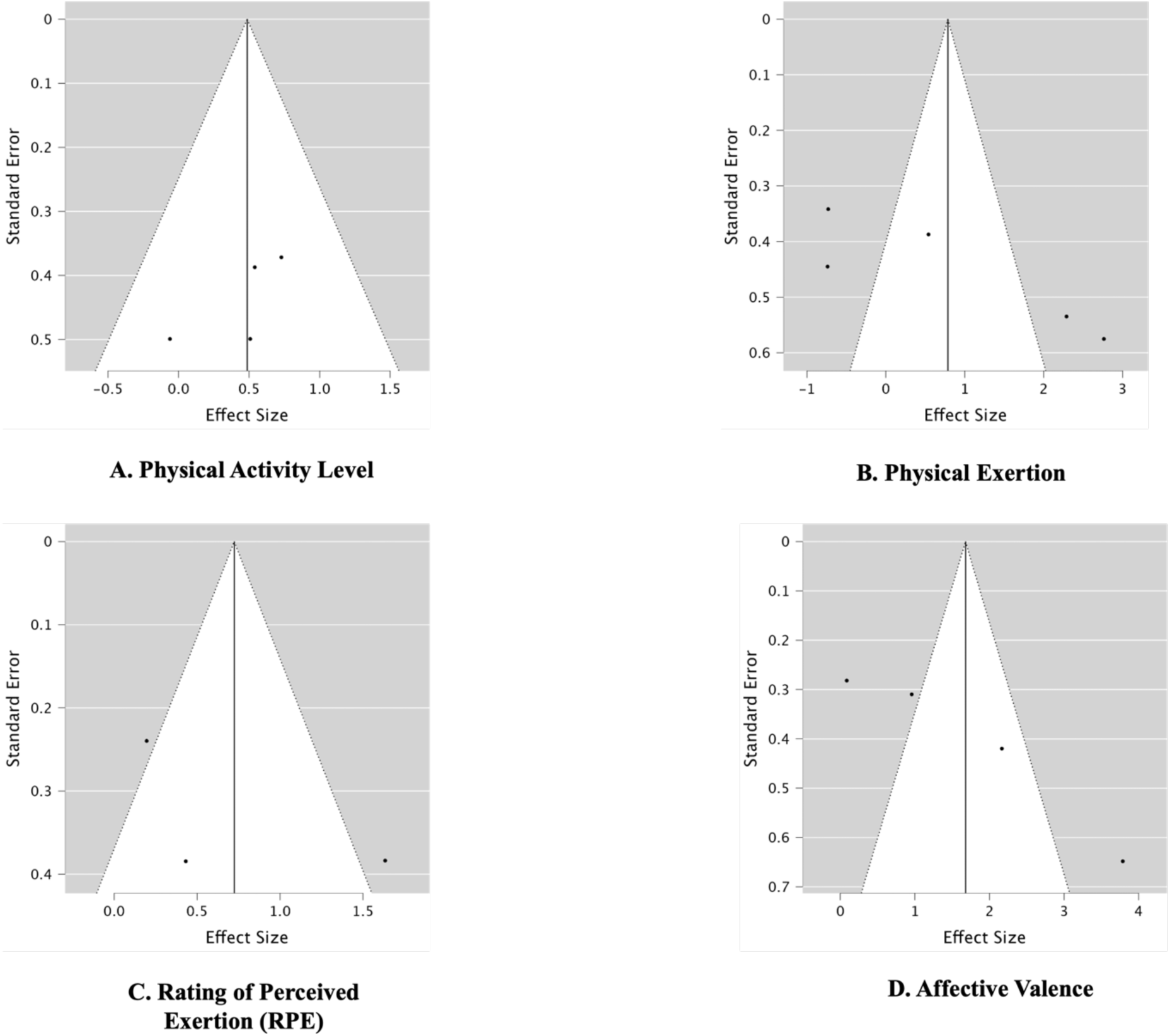
Funnel plots for Physical Activity Level (A), Physical Exertion (B), Rating of Perceived Exertion (RPE) (C), and Affective Valence (D).

## Discussion

The aim of this review was to systematically evaluate the effectiveness of Personalised Interactive Music Systems (PIMS) across physical activity levels, physiological outcomes (e.g., heart rate and step frequency), psychophysical outcomes (e.g., the rating of perceived exertion), and affective valence in relation to physical activity and exercise behaviours. A central focus was an exploratory meta-analysis of PIMS across these outcome domains.

The exploratory meta-analysis revealed that PIMS demonstrate favourable effects on physical activity levels and affective valence, with effect size estimates surpassing those of general music listening [6]. However, the certainty of evidence is limited by methodological inconsistencies, a moderate to high risk of bias and the limited number of published studies eligible for meta-analyses. Importantly, no significant effects were observed for ratings of perceived exertion (RPE) or measured physical exertion. This reflects variability in the psychophysical outcomes associated with interventions using PIMS.

When examining the findings of individual studies separately, they offer preliminary evidence that PIMS may improve physical, psychophysical, and affective outcomes related to physical activity and exercise. For example, [22] observed longer exercise durations during sessions utilizing Jymmin® compared to routines with passive music listening, without significant increases in perceived exertion. Similarly, [50] reported increased weekly physical activity volumes among cardiovascular disease patients using personalised Rhythmic Auditory Stimulation (RAS)-enhanced playlists. Additionally, [60] provided qualitative evidence suggesting that PIMS may prompt physical activity, such as reducing sitting time in office settings.

However, interpreting these findings is challenging due to methodological limitations and variability in population characteristics. Some studies focus on clinical populations, such as cardiovascular disease patients [50], while others target healthy younger adults [53] or elderly participants [22]. Several studies lack demographic details entirely, further complicating the assessment of population-specific efficacy. Sample sizes also vary widely, from single participants [51] to larger groups [53, *N* = 36].

### Heterogeneity in PIMS outcomes: methodological influences

The wide prediction intervals observed across outcome domains reflect the substantial heterogeneity in PIMS effects. For example, prediction intervals for physical activity levels and affective valence highlight significant variability in potential effect sizes. This suggests that while PIMS may provide positive average effects, individual study outcomes could range from substantial benefits to negligible or even negative impacts. Similarly, the prediction intervals for RPE and physical exertion emphasise uncertainty surrounding these psychophysical outcomes, pointing to inconsistencies in measurement and intervention design.

Specifically, variations in study methodologies and control group conditions contribute significantly to this heterogeneity. Some studies utilised passive music or other auditory stimuli as controls, while others used no-music conditions. This negatively affects comparability. Well-powered randomised designs, such as [50], produced robust findings, whereas smaller studies, such as [22], yielded non-significant results, pointing to the influence of study design and statistical power. Additionally, short intervention durations and small sample sizes [55,57] constrain the generalisability of findings. The absence of standardised metrics and protocols across studies further hinders the ability to synthesise outcomes and develop systematic guidelines for PIMS interventions. To alleviate this, future research should adopt standardised protocols and outcome measures. This could be achieved via a Music Selection and Delivery Protocol, ensuring uniformity through a predefined library of music tracks categorised by tempo and intensity, delivered via standardised systems (e.g., wireless headphones at consistent volumes). Validated tools such as the Borg Rating of Perceived Exertion (RPE) and the Feeling Scale (FS) for measuring affective valence, administered at fixed intervals, may enhance comparability.

### Feasibility of PIMS on physical activity levels and affective outcomes

Despite methodological inconsistencies, our findings suggest that PIMS may have a positive influence on physical activity levels. Studies in this cluster were rated as having low [50] to moderate [12,22] risk of bias, with both [50] and [22] focusing on elderly populations.

Positive effects include increased exercise duration [22; ∼66 seconds] and overall weekly physical activity [50; ∼105.4 additional minutes per week on average]. However, [12] found no significant impact of PIMS on physical activity levels. The low heterogeneity in this cluster indicates consistent findings despite variations in study design and participant populations. This is promising and calls for further investigation.

Our results align with Clark et al. (2024), who noted that music listening, when combined with physical activity, enhances exercise outcomes in older adults. Both [50] and [12] employed synchronisation strategies—RAS and auditory-motor coupling, respectively— consistent with frameworks by [8,67] that link synchronised music to improved physical activity and exercise performance. However, the exploratory nature of the meta-analysis and the small number of studies limit the potential generalisability of these findings. Further research with diverse populations and robust methodologies is required to confirm whether PIMS are effective adjuncts for increasing physical activity levels.

For affective valence, the large effect size estimate suggests PIMS contribute to elevated affective experiences during physical activity and exercise [12,22,34,52]. However, this finding is strongly influenced by [12], whose notably high effect size estimate substantially raised the overall meta-analytic effect size estimate. In contrast, smaller effects observed in other studies [22,34] reduced the precision and generalisability of the overall meta-analytic finding. The differences in these outcomes likely reflect variations in music selection methods: researcher-selected music in [12] prompted synchronisation and enjoyment (‘fun and enjoyment’ ratings via IMI), while self-selected music [52] and device-generated feedback [22,34] influenced affective outcomes in distinct ways. In [12] researcher-selected music facilitated synchronisation, while [52] used self-selected music based on participants’ individual preferences. [22,34] utilised device-generated musical feedback, where participants’ movements influenced the music. These differences suggest that PIMS may enhance affective valence outcomes during physical activity and exercise through both self-selected and researcher-selected music, with evidence of positive effects for music tailored to individual preferences (aligning with prior research by [7,11]) as well as for standardised, researcher-selected stimuli.

Curiously, [12] reported no significant benefits for RPE, despite utilising auditory-motor coupling strategies. This discrepancy may find alignment with Dual-Mode Theory, as even though music can enhance automatic synchronisation and facilitate improved physical performance, it does not always mitigate RPE if reflective processes (e.g., cognitive appraisal of effort) are less engaged [13]. The substantial heterogeneity within the affective valence cluster, driven by variability in musical strategies, participant demographics, and inconsistent measurement tools (e.g., MDMQ, IMI, FS), further supports Affective-Reflective Theory’s (ART) assertion that individual and contextual factors critically shape affective outcomes during exercise.

All studies in the affective valence cluster were deemed to have a moderate risk of bias. Furthermore, the reliance on measurement scales without strong theoretical grounding, as noted in [12], suggests the need for alignment with validated frameworks such as ART. For instance, the Feeling Scale (FS) used by [52] directly measures the pleasure-displeasure dimensions central to ART, aligning with validated frameworks in physical activity and exercise contexts [68]. The FS provides a theoretically robust and context-specific assessment of affective responses, capturing the transient emotional states during exercise that ART posits are critical for shaping future behavioural intentions. These findings tentatively indicate that these PIMS leverage momentary affective responses to improve exercise experiences [6,7,17]. In sum, findings across the physical activity and affective valence meta-analytic clusters suggest PIMS may support affect augmentation during physical activity, highlighting their potential to enhance both physical activity levels and affective outcomes [5,17].

### PIMS tempo adjustments and synchronisation in physical activity and exercise outcomes

The identification of faster music tempi as a statistically significant moderator in the meta-regression aligns with evidence supporting the role of synchronisation strength and auditory-motor coupling in enhancing exercise outcomes [8,45]. For instance, faster tempi provide consistent rhythmic cues that facilitate the alignment of motor actions with auditory stimuli. This can optimise auditory-motor coupling [8–10] which, in turn, enables predictive synchronisation to reduce RPE [7]. For example, [52] reported that real-time tempo adjustments based on heart rate significantly reduced RPE and improved affective responses. This indicates that synchronised music facilitated participants’ dissociation from internal sensory signals and promoted enjoyment during exercise [7].

### Limitations and future directions

This review presents the first systematic exploration of PIMS exclusively within physical, psychophysical, and affective domains of physical activity and exercise. While it provides valuable insights, several limitations must be acknowledged. A significant proportion of the included studies (14 out of 18) primarily assessed the feasibility of PIMS, with few investigating direct outcomes related to physical activity or exercise. Many experimental studies were limited by short durations, small sample sizes, and insufficiently rigorous methodologies. Similarly, proof-of-concept and user-testing studies largely focused on system feasibility rather than assessing objective psychophysiological outcomes.

Consequently, the high risk of bias in 10 studies underscores the overall low quality of evidence. Additionally, the small number of eligible studies precluded sensitivity analyses which further emphasises the preliminary nature of this review’s findings.

Few studies identified physical activity as a primary outcome, often relegating it to secondary importance. Objective assessments of physical activity—such as measures of frequency, intensity, and duration—were notably absent, making it difficult to draw robust conclusions or compare results across studies. Standardising methods for quantifying physical activity would enhance future research by enabling more meaningful cross-study comparisons.

Furthermore, the methodology used in this study was limited by substantial heterogeneity across studies. This prevents a unified meta-analysis and necessitates the reporting of separate outcomes. Variability in study designs, participant demographics, and measurement tools contributed to unexplained heterogeneity, while the small number of studies precluded sensitivity analyses. These factors, combined with the exploratory nature of the meta-analysis, point to the need for standardised methodologies and rigorous reporting in future research.

To address these limitations, future research should prioritise larger, randomised controlled trials with diverse populations and longer intervention periods. Longitudinal studies are particularly needed to evaluate the sustained impact of PIMS on physical activity and exercise. Additionally, investigating the mechanisms underlying individual variability in PIMS responses could optimise these systems for different populations and exercise contexts. This highlights the need for more rigorous research to validate these effects and refine PIMS interventions, particularly through the development of dynamic systems that can adapt tempo in real time to suit diverse user needs and exercise contexts [59,67].

Emerging trends in PIMS, such as music recommender systems examined by [47–49], highlight the potential for integration with streaming services such as Spotify. These systems demonstrated promising user feedback [48] and feasibility, suggesting they could serve as a foundation for future hypothesis-driven studies. Incorporating feedback from wearable and smartphone devices offers another avenue for development, allowing PIMS to adapt based on physical activity and exercise metrics as well as music preferences. Finally, many PIMS are relatively low-cost interventions (e.g., the devices in the [50] study cost approximately $75 per patient) and could have significant cost-effectiveness implications as part of broader health policy strategies to enhance physical activity and exercise participation at the population level [5].

## Conclusions

This systematic review provides exploratory evidence that PIMS may positively impact physical activity levels and affective valence in physical activity and exercise contexts. The meta-analysis revealed moderate effect sizes for physical activity levels and significant but heterogeneously distributed effects for affective valence. However, outcomes for RPE and physical exertion were inconclusive due to high heterogeneity and limited study quality.

The findings are constrained by methodological limitations, including high risk of bias, small sample sizes, short study durations, and inconsistent measures across studies. Furthermore, the lack of theoretical frameworks for informing PIMS designs and the absence of standardisation in quantifying physical activity outcomes limit the generalisability of these findings. PIMS remain considerably underexplored, and further research is essential.

Overall, PIMS provide promising potential for enhancing physical activity levels and elevated affective valence, offering engaging physical activity and exercise opportunities for the public at large. With advancements in adaptive systems capable of real-time tempo adjustments, PIMS may emerge as effective adjuncts for physical activity and exercise, pending rigorous validation in diverse populations.

## Data Availability

The authors confirm that the data supporting the findings of this study are available within the article [and/or] its supplementary materials accessed here: OSF, https://osf.io/jpy5k/

## Author Contributions

**Conceptualisation:** Andrew Danso, Tiia Kekäläinen, Geoff Luck, Sebastien Chastin

**Methodology:** Andrew Danso, Tiia Kekäläinen, Geoff Luck

**Investigation:** Andrew Danso, Tiia Kekäläinen, Iballa Burunat, Pedro Neto, Anastasios Mavrolampados, William M. Randall

**Data Curation:** Andrew Danso, Tiia Kekäläinen, Patti Nijhuis, Iballa Burunat, Pedro Neto, Anastasios Mavrolampados, William M. Randall

**Formal Analysis:** Andrew Danso, Keegan Knittle

**Writing - Original Draft:** Andrew Danso, Friederike Koehler, Patti Nijhuis, Rebekah Rousi, Kat Agres, Jennifer MacRitchie, Niels Chr. Hansen

**Writing - Review & Editing:** Andrew Danso, Tiia Kekäläinen, Friederike Koehler, Keegan Knittle, Patti Nijhuis, Iballa Burunat, Pedro Neto, Anastasios Mavrolampados, William M. Randall, Niels Chr. Hansen, Alessandro Ansani, Timo Rantalainen, Vinoo Alluri, Martin Hartmann, Rebecca S. Schaefer, Johanna Ihalainen, Rebekah Rousi, Kat Agres, Jennifer MacRitchie, Petri Toiviainen, Suvi Saarikallio, Sebastien Chastin, Geoff Luck

## Supporting information

### S1 Appendix. Operationalisation of Terms

- **Physical activity level** encompasses the volume, intensity, and compliance with physical activity recommendations or exercise regimens. Volume represents the overall amount of physical activity, and can be measured using metrics such as mean total counts per day [29]. Intensity can be categorised as absolute intensity using metabolic equivalents (METs) or relative intensity expressed as a percentage of an individual’s maximum oxygen uptake reserve (%VO2R, [30]). Compliance, referring to adherence to recommendations or regimens, can be assessed by tracking changes in physical activity volume, monitoring device usage, or measuring adherence to target heart-rate zones during physical activity or acute bouts of physical exercise [29].
- **Affective valence** refers to the emotional response of pleasure or displeasure experienced during or after physical activity. It focuses specifically on the valence dimension of emotional states—how positive or negative the feeling is—without incorporating arousal (activation). Affective valence captures moment-to-moment changes in an individual’s subjective emotional state as they engage in physical activity. For instance, physical activity often elicits feelings of pleasure (positive affect), though the intensity and context of the activity, as well as individual differences, influence these responses.
- **Rating of perceived exertion (RPE)** is a subjective numerical value that individuals assign to their sense of how hard their body is working during physical activity [7,37]. It is formed from a multitude of sensory cues, integrating both physiological sensations and psychological perceptions [37]. Sensations from muscles, skin, and joints, and effects stemming from the cardio-pulmonary system all contribute to this overall perception. The Borg RPE scale is specifically designed to measure perceived exertion during steady-state aerobic exercise, such as cycling or running. The commonly used Borg CR-10 scale, is a category-ratio scale, designed to measure the perceived intensity of various sensations, experiences, and feelings. Its primary application is in assessing perceived exertion but can also gauge other subjective experiences.
- **Physical exertion** is the effort exerted by the body to perform physical activity, characterised by the physiological, biomechanical, and perceptual demands placed on the individual [35]. Heart rate is a fundamental physiological indicator of exertion, reflecting the cardiovascular system’s response to physical demands [35]. Stride length and pace are essential biomechanical parameters, particularly in activities such as running and walking [35].

### S2 Appendix. Description of the outcome measures used in the studies

- **Physical Activity Level:** [50] measured physical activity level by mean weekly minutes of physical activity, which was captured using a tri-axial accelerometer worn by the participants. [22] measured physical activity level through the duration of exercise until exhaustion, timed with a stopwatch. [12] measured compliance with a prescribed exercise regime by monitoring participants’ adherence to target heart rate zones during a cycling session.
- **Affective valence:** [52] measured affective valence using the Feeling Scale (FS). The FS is based on Russell’s circumplex model of affect [14,33]. The FS assesses affective valence (how positive or negative someone feels). [22,34] used the Multidimensional Mood Questionnaire (MDMQ) [32] to study the effects of PIMS on mood during acute bouts of physical exercise. The MDMQ includes subscales for “good vs. bad mood,” ”calmness vs. agitation,” and “alertness vs. tiredness.” While the MDMQ’s three subscales generally correspond to the circumplex model of affect [14,33] only the “good vs. bad mood” subscale was used, as it specifically aligns with the pleasure-displeasure (affective valence) dimension of affect. [12] examined the effect of PIMS on intrinsic motivation using an “interest/enjoyment” subscale of the Intrinsic Motivation Inventory (IMI) [62] during acute bouts of physical exercise. While not a direct measurement of affect, this subscale reflects positive valence and enjoyment, aligning broadly with the pleasure (affective valence) dimension of affect [63].
- **RPE:** All three studies that assessed Rating of Perceived Exertion (RPE) used the Borg CR-10 scale to capture participants’ subjective experience of exercise intensity. [22,52] both employed the scale, with [52] collecting RPE ratings at specific time intervals during the exercise, while [22] opted for more frequent measurements every 90 seconds and at the conclusion of the exercise. [12] also utilised the Borg CR-10 scale, though the precise timing of their measurements is not detailed.
- **Physical Exertion:**[52] measured heart rate as a physiological measure of exertion using a Polar Verity Sense upper-arm heart-rate measurement device based on photoplethysmography (PPG). [54] measured pace by calculating Swings Per Minute (SWPM) using data from the accelerometer on a smartphone. [12] measured heart rate via a Polar T61 heart rate belt.

**S1 Table.** Descriptions and outcomes of the PIMS used in the nine experimental studies.

| Reference | PIMS Used | PIMS Description | Outcomes |
| --- | --- | --- | --- |
| [50] | Music audio-playlists | Personalised music audio-playlists to improve adherence to physical activity among cardiovascular disease patients participating in a structured exercise program. | Control (No Music), 370.2 minutes, $p < .001^a$<br>Music Playlists, 475.6 minutes, $p < .001^a$<br>Music Playlists with RAS, 631.3 minutes |
| [53] | Music Assisted Run Trainer (MART) | A smartphone application, to assist jogging activity by adapting music tempo based on the user's step frequency or heart rate. | Aligning music tempo with step frequency – targeting optimal heart rate for cardiovascular training. |
| [48] | Music Recommendation System | A music recommendation system to motivate individuals to exercise more effectively by incorporating user profiling and reinforcement learning. | Improved satisfaction with playlists, reduced number of rejections needed to finalise a playlist. |
| [34] | Jymmin® | Exercise with musical feedback | Significant main effect of the "jymmin" condition on mood enhancement vs. passive listening, with a reported $F(1, 43) = 10.67$, $p < .05$ |
| [56] | SoundBike | A stationary bicycle system designed to enhance cyclists' spontaneous synchronisation with external music through musical sonification | Sonification using a beep at the point of maximum pedal pressure significantly increased cyclists' synchronisation strength with external music compared to no sonification ($p < .05$) and improved pedal cadence stability ($p < .05$). The beep condition resulted in a medium to large effect size for synchronisation and stability compared to other conditions. |
| [59] | D-Jogger | An adaptive music player designed to align music with the user's walking or running pace. | Alignment Strategy 1 – Minor increase in steps synchronised with beat.<br>Alignment Strategy 2 – Minor increase in steps synchronised with beat.<br>Alignment Strategy 3 – Tempo adjusts to match runner's pace throughout. Increase in synchronised steps; high phase-lock stability.<br>Alignment Strategy 4 – Adjusts phase and tempo during song to match each beat to footfall. Increase in synchronised steps; highest phase-lock stability. |
| [22] | Jymmin® | Exercise with musical feedback | Participants exercised significantly longer with Jymmin® (Jymmin® workout 248.75 seconds vs. Conventional workout 182.73 seconds) |
| [60] | Flow Platform (Smart Cushion) | A flow platform (a smart cushion) which used interactive music to cue office workers to reduce their sedentary time. | Both interactive and continuous music were similarly effective in motivating posture changes and reducing sedentary behaviour. |
| [12] | MoBeat | Intensity-based coaching during exercise by giving real-time feedback on training pace and intensity through interactive music. | The moBeat system was found to have a significant positive effect on intrinsic motivation ($p < .001$) and attentional focus ($p < .001$) during exercise. The moBeat system did not significantly reduce perceived exertion ($p = .266$) compared to the reference system. |

<sup>a</sup>= $p$ -value compares music playlists vs. a control group with no music. The $p$ -value for the RAS-enhanced group compared to the others is not specified here but indicated as significant in the study.

**S2 Table.** Descriptions and outcomes of the PIMS used in the nine Proof of Concept and User Testing Studies.

| Reference | PIMS Used | PIMS Description | Outcomes |
| --- | --- | --- | --- |
| [47] | The DJ Running System | The system dynamically adapts music using sensor data from physical movements or exercise equipment. | N/A |
| [51] | Music Feedback Exercise (MFE) | The study integrates MFE into Soundjack, using sensors to modulate music playback based on exercise intensity. | Positive feedback on system responsiveness and music integration. |
| [52] | Music-Assisted Internet of Things (IoT) Exercise System | The system adapts music tempo in real-time to the runner's heart rate. | N/A |
| [54] | Runner's Jukebox (RJ) | RJ uses a pace recognition algorithm enabling the music player to play songs matched with the user pace and adjust playback speed dynamically to follow the user's pace changes. | Fixed BPM and pace matching gave better exercise effect (SWPM) vs. no music and vs. randomly selected music |
| [55] | A Stationary bike augmented in an audio reality environment. | The use of sensors to monitor the user's pace and heart rate while exercising on a stationary bike. Audio feedback and cues are manipulated based on the user's performance. | N/A |
| [58] | DJogger | A music interface to leverage body movement in order to select music and adapt its tempo to the user's pace, focusing on entrainment – synchronisation between music and walking. | The majority of subjects synchronised to the beats, no matter what the users' pace.<br>If the music tempo is close enough to the user's pace, the user tends to synchronise his/her steps with the beats. |
| [57] | Sonification of accelerometry data (recordings of physical activity) | This system transforms daily physical activity data recorded by wearable devices into musical pieces. | The majority of participants correctly identified the musical sonification associated with physical activity. |
| [49] | Context-Aware Music Recommender System (CAMRS) | The system classified physical activities by analysing data from the smartphone's accelerometer, allowing it to predict the user's activity. It recommends music with a Beats Per Minute (BPM) value to match the intensity of the detected physical activity. | When using the system, rates of perceived effort decreased in the majority of cases, and mood improved in the majority of cases. |
| [61] | Interactive Music System for Alzheimer's patients | This music system for Alzheimer's patients, focuses on how dynamically adapting musical beats and rhythms can stimulate and motivate physical activity. The system adapts the tempo of music to match the pace of repetitive bodily movements. | The interactive music system produces an entraining effect on participants. |

## References

1 Bull FC, Al-Ansari SS, Biddle S, Borodulin K, Buman MP, Cardon G, et al. World Health Organization 2020 guidelines on physical activity and sedentary behaviour. Br J Sports Med. 2020;54: 1451–1462. doi:10.1136/bjsports-2020-102955

1. Posadzki P, Pieper D, Bajpai R, Makaruk H, Könsgen N, Neuhaus AL, et al. Exercise/physical activity and health outcomes: an overview of Cochrane systematic reviews. BMC Public Health. 2020;20: 1724–1724. doi:10.1186/s12889-020-09855-3

2. Caspersen CJ, Powell KE, Christenson GM. Physical Activity, Exercise, and Physical Fitness: Definitions and Distinctions for Health-Related Research. Public Health Rep 1974-.1985;100: 126–131.

3. Chase JAD. Systematic review of physical activity intervention studies after cardiac rehabilitation. J Cardiovasc Nurs. 2011;26: 351–358. doi:10.1097/JCN.0b013e3182049f00

4. Moy ML, Teylan M, Weston NA, Gagnon DR, Garshick E. Daily Step Count Predicts Acute Exacerbations in a US Cohort with COPD. PLoS ONE. 2013;8. doi:10.1371/journal.pone.0060400

5. Clark IN, Taylor NF, Peiris CL. Music listening interventions for physical activity: a systematic review and meta-analysis of randomised controlled trials. Disabil Rehabil. 2024;46: 13–20. doi:10.1080/09638288.2022.2155715

6. Terry PC, Karageorghis CI, Curran ML, Martin OV, Parsons-Smith RL. Effects of music in exercise and sport: A meta-analytic review. Psychol Bull. 2020;146: 91–117. doi:10.1037/bul0000216

7. Bood RJ, Nijssen M, van der Kamp J, Roerdink M. The Power of Auditory-Motor Synchronization in Sports: Enhancing Running Performance by Coupling Cadence with the Right Beats. PLoS ONE. 2013;8. doi:10.1371/journal.pone.0070758

8. Karageorghis CI, Priest D-L. Music in the exercise domain: a review and synthesis (Part I). Int Rev Sport Exerc Psychol. 2012;5: 44–66. doi:10.1080/1750984X.2011.631026

9. Edworthy J, Waring H. The effects of music tempo and loudness level on treadmill exercise. Ergonomics. 2006;49: 1597–1610. doi:10.1080/00140130600899104

10. Khalfa S, Roy M, Rainville P, Dalla Bella S, Peretz I. Role of tempo entrainment in psychophysiological differentiation of happy and sad music? Int J Psychophysiol. 2008;68: 17–26. doi:10.1016/j.ijpsycho.2007.12.001

11. Van Der Vlist B, Bartneck C, Mäueler S. moBeat: Using interactive music to guide and motivate users during aerobic exercising. Appl Psychophysiol Biofeedback. 2011;36: 135–145. doi:10.1007/s10484-011-9149-y

12. Ekkekakis P. Pleasure and displeasure from the body: Perspectives from exercise. Cogn Emot. 2003;17: 213–239. doi:10.1080/02699930302292

13. Russell JA. Core affect and the psychological construction of emotion. Psychol Rev. 2003;110: 145–172. doi:10.1037/0033-295X.110.1.145

14. Brand R, Ekkekakis P. Affective–Reflective Theory of physical inactivity and exercise. Ger J Exerc Sport Res. 2018;48: 48–58. doi:10.1007/s12662-017-0477-9

15. Karageorghis CI. The scientific application of music in exercise and sport: Towards a new theoretical model. Sport Exerc Psychol 2nd Ed. 2016; 276–322.

16. Karageorghis CI. Music-Related Interventions in the Exercise Domain. Handbook of Sport Psychology. Wiley; 2020. pp. 929–949. doi:10.1002/9781119568124.ch45

17. Agres KR, Schaefer RS, Volk A, van Hooren S, Holzapfel A, Dalla Bella S, et al. Music, Computing, and Health: A Roadmap for the Current and Future Roles of Music Technology for Health Care and Well-Being. Music Sci. 2021;4: 205920432199770– 205920432199770. doi:10.1177/2059204321997709

19. Danso A. The Use of Technology in Music-based Interventions for Health and Education. Dissertation, University of Jyväskylä. 2023.

19. Marcus B, Owen N, Forsyth L, Cavill N, Fridinger F. Physical activity interventions using mass media, print media, and information technology. Am J Prev Med. 1998;15: 362–378. doi:10.1016/S0749-3797(98)00079-8

20. Wijnalda G, Pauws S, Vignoli F, Stuckenschmidt H. A personalized music system for motivation in sport performance. IEEE Pervasive Comput. 2005;4: 26–32. doi:10.1109/MPRV.2005.47

22. Rehfeld K, Hans Fritz T, Prinz A, Schneider L, Villringer A, Witte K. Musical feedback system Jymmin ® leads to enhanced physical endurance in the elderly-A feasibility study. 2022.

22. Laranjo L, Ding D, Heleno B, Kocaballi B, Quiroz JC, Tong HL, et al. Do smartphone applications and activity trackers increase physical activity in adults? Systematic review, meta-analysis and metaregression. Br J Sports Med. 2021;55: 422–432. doi:10.1136/bjsports-2020-102892

23. Michie S, Richardson M, Johnston M, Abraham C, Francis J, Hardeman W, et al. The Behavior Change Technique Taxonomy (v1) of 93 Hierarchically Clustered Techniques: Building an International Consensus for the Reporting of Behavior Change Interventions. Ann Behav Med. 2013;46: 81–95. doi:10.1007/s12160-013-9486-6

24. Ziv G, Lidor R. Music, Exercise Performance, and Adherence in Clinical Populations and in the Elderly: A Review. J Clin Sport Psychol. 2011;5: 1–23. doi:10.1123/jcsp.5.1.1

25. Page MJ, McKenzie JE, Bossuyt PM, Boutron I, Hoffmann TC, Mulrow CD, et al. The PRISMA 2020 statement: an updated guideline for reporting systematic reviews. BMJ. 2021; n71–n71. doi:10.1136/bmj.n71

26. van de Schoot R, de Bruin J, Schram R, Zahedi P, de Boer J, Weijdema F, et al. An open source machine learning framework for efficient and transparent systematic reviews. Nat Mach Intell. 2021;3: 125–133. doi:10.1038/s42256-020-00287-7

27. Ouzzani M, Hammady H, Fedorowicz Z, Elmagarmid A. Rayyan—a web and mobile app for systematic reviews. Syst Rev. 2016;5: 210–210. doi:10.1186/s13643-016-0384-4

28. Bassett DR, Troiano RP, Mcclain JJ, Wolff DL. Accelerometer-based Physical Activity: Total Volume per Day and Standardized Measures. Med Sci Sports Exerc. 2015;47: 833–838. doi:10.1249/MSS.0000000000000468

29. Kujala UM, Pietilä J, Myllymäki T, Mutikainen S, Föhr T, Korhonen I, et al. Physical Activity: Absolute Intensity versus Relative-to-Fitness-Level Volumes. Med Sci Sports Exerc. 2017;49:<otherinfo> 4</otherinfo>74–481. doi:10.1249/MSS.0000000000001134

30. Hardy CJ, Rejeski WJ. Not What, but How One Feels: The Measurement of Affect during Exercise. J Sport Exerc Psychol. 1989;11: 304–317. doi:10.1123/jsep.11.3.304

31. Steyer R, Schwenkmezger P, Notz P, Eid M. Development of the Multidimensional Mood State Questionnaire (MDBF). Primary dataEntwicklung des Mehrdimensionalen Befindlichkeitsfragebogens (MDBF). Primärdatensatz. ZPID Leibniz Institute for Psychology; 2003. doi:10.5160/PSYCHDATA.SRRF91EN15

32. Ekkekakis P, Petruzzello SJ. Analysis of the affect measurement conundrum in exercise psychology. Psychol Sport Exerc. 2000;1: 71–88. doi:10.1016/S1469-0292(00)00010-8

33. Fritz TH, Halfpaap J, Grahl S, Kirkland A, Villringer A. Musical feedback during exercise machine workout enhances mood. Front Psychol. 2013;4. doi:10.3389/fpsyg.2013.00921

34. Nystoriak MA, Bhatnagar A. Cardiovascular Effects and Benefits of Exercise. Front Cardiovasc Med. 2018;5: 135. doi:10.3389/fcvm.2018.00135

35. Borg G. Perceived exertion as an indicator of somatic stress. Scand J Rehabil Med. 1970;2: 92–98.

36. Borg G. Psychophysical bases of perceived exertion. Med Sci Sports Exerc. 1982;14: 377–381.

37. Grammatopoulos T, Hunter JWS, Munn Z, Stone JC, Barker TH. Reporting quality and risk of bias in JBI systematic reviews evaluating the effectiveness of interventions: a methodological review protocol. JBI Evid Synth. 2023;21: 584–591. doi:10.11124/JBIES-22-00317

38. Campbell M, McKenzie JE, Sowden A, Katikireddi SV, Brennan SE, Ellis S, et al. Synthesis without meta-analysis (SWiM) in systematic reviews: reporting guideline. BMJ. 2020; l6890. doi:10.1136/bmj.l6890

39. Wilson, D. Practical Meta-Analysis Effect Size Calculator. In: Practical Meta-Analysis Effect Size Calculator [Online calculator, version 2023.11.27] [Internet]. 27 Nov 2023 [cited 4 Apr 2024]. Available: https://campbellcollaboration.org/research-resources/effect-size-calculator.html

40. Bender R, Friede T, Koch A, Kuss O, Schlattmann P, Schwarzer G, et al. Methods for evidence synthesis in the case of very few studies. Res Synth Methods. 2018;9: 382–392. doi:10.1002/jrsm.1297

41. Borenstein M. How to understand and report heterogeneity in a meta-analysis: The difference between I-squared and prediction intervals. Integr Med Res. 2023;12: 101014. doi:10.1016/j.imr.2023.101014

42. Sterne JAC, Egger M. Funnel plots for detecting bias in meta-analysis. J Clin Epidemiol. 2001;54: 1046–1055. doi:10.1016/S0895-4356(01)00377-8

43. Sterne JAC, Harbord RM. Funnel Plots in Meta-analysis. Stata J Promot Commun Stat Stata. 2004;4: 127–141. doi:10.1177/1536867X0400400204

44. Elliott D. Music During Exercise: Does Tempo Influence Psychophysical Responses? 2007.

45. Jetté M, Sidney K, Blümchen G. Metabolic equivalents (METS) in exercise testing, exercise prescription, and evaluation of functional capacity. Clin Cardiol. 1990;13: 555–565. doi:10.1002/clc.4960130809

46. Álvarez P, Zarazaga-Soria FJ, Baldassarri S. Mobile music recommendations for runners based on location and emotions: The DJ-Running system. Pervasive Mob Comput. 2020;67. doi:10.1016/j.pmcj.2020.101242

47. Fang J, Grunberg D, Lui S, Wang Y. Development of a music recommendation system for motivating exercise. Institute of Electrical and Electronics Engineers Inc.; 2018. pp. 83–86. doi:10.1109/ICOT.2017.8336094

48. Ospina-Bohórquez A, Gil-González AB, Moreno-García MN, de Luis-Reboredo A. Context-aware music recommender system based on automatic detection of the user’s physical activity. Springer; 2021. pp. 142–151. doi:10.1007/978-3-030-53036-5_15

49. Alter DA, O’Sullivan M, Oh PI, Redelmeier DA, Marzolini S, Liu R, et al. Synchronized personalized music audio-playlists to improve adherence to physical activity among patients participating in a structured exercise program: a proof-of-principle feasibility study. Sports Med - Open. 2015;1. doi:10.1186/s40798-015-0017-9

50. Carôt A, Fritz TH, Englert K. Towards a System Supporting Music Feedback Exercise in Physical Tele-Rehabilitation. Institute of Electrical and Electronics Engineers (IEEE); 2023. pp. 1–7. doi:10.1109/ieeeconf59510.2023.10335285

51. Chen Y, Chen CC, Tang LC, Chieng WH. Mid-Distance Running Exercise Assistance System via IoT and Exercise-oriented Music. Institute of Electrical and Electronics Engineers Inc.; 2023. pp. 264–272. doi:10.1109/AIIoT58121.2023.10174324

53. Chen L-Y, Tung Y-J, Jang J-SR. MART-Music Assisted Run Trainer. 2014.

53. Jun S, Rew J, Hwang E. Runner’s Jukebox: A Music Player for Running Using Pace Recognition and Music Mixing. 2015.

54. Maculewicz J, Serafin S. A Stationary Bike in Augmented Audio Reality. Association for Computing Machinery; 2015. pp. 164–167. doi:10.1145/2838944.2838984

55. Maes P-J, Lorenzoni V, Six J. The SoundBike: musical sonification strategies to enhance cyclists’ spontaneous synchronization to external music. J Multimodal User Interfaces. 2019;13: 155–166. doi:10.1007/s12193-018-0279-x

56. Mendoza JI, Danso A, Luck G, Rantalainen T, Palmberg L, Chastin S. Musification of Accelerometry Data Towards Raising Awareness of Physical Activity. 2022 [cited 16 Dec 2024]. doi:10.5281/ZENODO.7243875

58. Moens Leon van Noorden Marc Leman IPEM B. D-JOGGER: SYNCING MUSIC WITH WALKING. Universidad Pompeu Fabra.; 2010. pp. 451–456. Available: http://www.yamaha.com/bodibeat/

58. Moens B, Muller C, Van Noorden L, Franěk M, Celie B, Boone J, et al. Encouraging spontaneous synchronisation with D-jogger, an adaptive music player that aligns movement and music. PLoS ONE. 2014;9. doi:10.1371/journal.pone.0114234

59. Ren X, Lu Y, Visser V, Le PDH, Van Den Burg R. Interaction matters? Exploring interactive music as a reminder to break sedentary office time. International Association for Automation and Robotics in Construction I.A.A.R.C); 2017. pp. 1099–1106. doi:10.22260/isarc2017/0151

60. Rosseland RB. Rune B. Rosseland Design and evaluation of an interactive music system for exercise and physical activity with Alzheimer’s patients. 2016 pp. 1904–500. Available: www.soundeffects.dk

61. Ryan RM, Mims V, Koestner R. Relation of reward contingency and interpersonal context to intrinsic motivation: A review and test using cognitive evaluation theory. J Pers Soc Psychol. 1983;45: 736–750. doi:10.1037/0022-3514.45.4.736

62. McAuley E, Duncan T, Tammen VV. Psychometric Properties of the Intrinsic Motivation Inventory in a Competitive Sport Setting: A Confirmatory Factor Analysis. Res Q Exerc Sport. 1989;60: 48–58. doi:10.1080/02701367.1989.10607413

63. McGuinness LA, Higgins JPT. Risk-of-bias VISualization (robvis): An R package and Shiny web app for visualizing risk-of-bias assessments. Res Synth Methods. 2021;12: 55–61. doi:10.1002/jrsm.1411

64. Cumpston M, Li T, Page MJ, Chandler J, Welch VA, Higgins JP, et al. Updated guidance for trusted systematic reviews: a new edition of the Cochrane Handbook for Systematic Reviews of Interventions. Cochrane Editorial Unit, editor. Cochrane Database Syst Rev. 2019 [cited 16 Dec 2024]. doi:10.1002/14651858.ED000142

65. Egger M, Smith GD, Schneider M, Minder C. Bias in meta-analysis detected by a simple, graphical test. bmj. 1997;315: 629–634.

66. Clark IN, Baker FA, Taylor NF. The modulating effects of music listening on health-related exercise and physical activity in adults: a systematic review and narrative synthesis. Nord J Music Ther. 2016;25: 76–104. doi:10.1080/08098131.2015.1008558

67. Bastos V, Rodrigues F, Davis P, Teixeira DS. Assessing affective valence and activation in resistance training with the feeling scale and the felt arousal scale: A systematic review. Kanawjia P, editor. PLOS ONE. 2023;18: e0294529. doi:10.1371/journal.pone.0294529

